# Comparison of AI-integrated pathways with human-AI interaction for population mammographic screening

**DOI:** 10.1101/2022.11.23.22282646

**Authors:** Helen ML Frazer, Carlos A Peña-Solorzano, Chun Fung Kwok, Michael S Elliott, Yuanhong Chen, Chong Wang, the BRAIx team, Jocelyn Lippey, John Hopper, Peter Brotchie, Gustavo Carneiro, Davis J McCarthy

## Abstract

Artificial intelligence (AI) holds promise for improving breast cancer screening, but many challenges remain in implementing AI tools in clinical screening services. AI readers compare favourably against individual human radiologists in detecting breast cancer in population screening programs. However, single AI or human readers cannot perform at the level of multi-reader systems such as those used in Australia, Sweden, the UK, and other countries. The implementation of AI readers in mammographic screening programs therefore demands integration of AI readers in multi-reader systems featuring collaboration between humans and AI. Successful integration of AI readers demands a better understanding of possible models of human-AI collaboration and exploration of the range of possible outcomes engendered by the effects on human readers of interacting with AI readers. Here, we used a large, high-quality retrospective mammography dataset from Victoria, Australia to conduct detailed simulations of five plausible AI-integrated screening pathways. We compared the performance of these AI-integrated pathways against the baseline standard-of-care “two reader plus third arbitration” system used in Australia. We examined the influence of positive, neutral, and negative human-AI interaction effects of varying strength to explore possibilities for upside, automation bias, and downside risk of human-AI collaboration. Replacing the second reader or allowing the AI reader to make high confidence decisions can improve upon the standard of care screening outcomes by 1.9–2.5% in sensitivity and up to 0.6% in specificity (with 4.6–10.9% reduction in the number of assessments and 48–80.7% reduction in the number of reads). Automation bias degrades performance in multi-reader settings but improves it for single-readers. Using an AI reader to triage between single and multi-reader pathways can improve performance given positive human-AI interaction. This study provides insight into feasible approaches for implementing human-AI collaboration in population mammographic screening, incorporating human-AI interaction effects. Our study provides evidence to support the urgent assessment of AI-integrated screening pathways with prospective studies to validate real-world performance and open routes to clinical adoption.

## Introduction

Breast cancer is the world’s most common cancer and a leading cause of cancer death in women [1]. Each year, approximately one million Australian women are screened under the BreastScreen Australia program [2], and there are challenges in accuracy, service experience, and time and cost efficiency. In 2020, approximately 59 per 10,000 participants were diagnosed with breast cancer and 16 per 10,000 participants were diagnosed with Ductal Carcinoma In Situ (DCIS) [2]. Despite a process of independent double reading of all mammograms by radiologists, and a third arbitration read when there is discordance (henceforth called “two reader with arbitration system”), in 2020 approximately 368 per 10,000 participants were recalled for assessment and later determined not to have breast cancer (false positive). Also, approximately 18.6 per 10,000 participants aged 50-74 years (2015-2017) subsequently discovered they had an “interval” breast cancer before their next scheduled screen after receiving an “all clear” result (false negative) [2].

Using artificial intelligence (AI) to help read mammograms has the potential to transform breast cancer screening by addressing the three key challenges of accuracy, client experience, and efficiency [2]. The evidence base for AI readers in breast cancer screening has been growing rapidly in recent years, with studies demonstrating the potential of AI to detect breast cancer on mammographic images with a similar accuracy to radiologists [3–11] and addressing key limitations of earlier concern [12]. Many of these studies evaluate the integration of AI into screening pathways via simulation, and they vary in the way human readers interact with the AI, where the AI is positioned in the screening pathway, and the specific screening pathway being simulated. This includes the complete replacement of current diagnosis pathways by an AI system [8, 13], using the AI as a decision referral system to diagnose low risk non-cancer cases without human intervention, while referring mid- and high-risk cases to human readers [14–16], or using a recall threshold to divide the cases into human review (mid-risk) and direct recall of (high-risk) suspicious cases [17]. Another option is to use AI in multiple positions in the pathway: first to rule out a percentage of the non-cancer cases without human intervention, then to rule in cases to supplemental imaging after being cleared by a double reading and consensus pathway [18]. Finally, in a double reading and consensus pathway, one of the human readers can be replaced by the AI to study the performance of the system before and after replacement [16, 19].

Previous studies have primarily focused on evaluating the feasibility and effectiveness of incorporating AI into existing screening pathways [5, 16, 17, 19–21]. These studies have shown promising results, often in terms of improved diagnostic accuracy and reduced workload, but they have been limited in the number of scenarios they evaluate [5, 17, 20] and typically avoid (statistically) simulating the arbitration read [19, 21] or any direct human-AI interaction [16]. Testing cohorts are not always representative or don’t have interval cancer follow-up and operating points are not always set on a separate dataset [12]. They are also primarily using commercial algorithms [16, 17, 19–21], which vary between studies and are assessed on different datasets, thereby precluding direct comparisons of the AI readers’ effectiveness as well as the viability of various AI integrated scenarios. There is a need for more in-depth analysis of where and how AI is best positioned within the screening pathway to maximise its benefits. This includes examining whether AI should be used as a primary screening tool, to assist radiologists in decision-making, or in a triage capacity to prioritize cases. An ongoing randomised controlled trial in Sweden [20], which has reported a positive interim safety milestone, has limited retrospective analysis of the reading pathway under test, in part due to its reliance on human-AI interaction. As human readers are likely to remain central to the decision making process, with concerns over the impacts of automation on radiologist performance over the short and long term [22], it is crucial to evaluate the potential impacts of human-AI interaction.

Here, we conducted detailed simulation studies incorporating human-AI interaction effects along with reader-level analysis of AI integrated screening scenarios using our retrospective testing cohort comprising data from over 90,000 screening clients in Victoria, Australia and over 600,000 mammogram images. Our simulations address limitations of previous work to gain insight into the performance of AI readers in collaboration with human readers and gain clinically-relevant guidance for the implementation of AI readers in screening programs. We used the BRAIx AI Reader (v3.0.7), a mammography classification model developed by the BRAIx research program based on an ensemble of open-source, image-based deep learning neural networks, trained on millions of screening mammograms and benchmarked on public datasets.

We analyse and compare five AI integrated screening pathways with the current standard of care, progressing from AI standalone, where AI has full autonomy in reading mammograms, to collaborative models like AI single-reader and AI reader-replacement, blending AI support with human expertise, and advanced integration methods like the AI band-pass and AI triage, which selectively engage AI in high-confidence tasks or pre-screening to direct workflow, aiming to increase screening accuracy and reduce the workload of human readers. The comparison of AI scenarios highlights how the effectiveness of AI integration varies across different screening pathways and roles. Uniquely, our simulation studies investigate possibilities for positive, neutral, and negative influence of AI integration on human reader performance for each of the five AI scenarios, deeply exploring the space of benefits and risks with AI integration.

We use our large, representative screening population alongside rich reader data to carefully simulate reader behaviour (especially that of the third reader in arbitration situations) and human-AI interaction to provide actionable information relevant to current directions for AI implementation [20], improve on previous efforts [14–16, 18, 19, 21], and offer insights into the optimal use of AI in enhancing screening outcomes.

## Results

### Study design

We evaluated the AI reader on a representative, population screening dataset collected from women who attended the BreastScreen Victoria program from 2016-2019 in Victoria, Australia. The screening program targets women aged 50-74 and typically collects four 2D mammograms for each client (left and right mediolateral oblique, MLO, and craniocaudal, CC) every two years. Each mammogram is read independently by two breast imaging radiologists and a third if there is disagreement (two reader with arbitration) who has access to the outputs of the first two readers. Readers flag clients for recall for further assessment if they detect indications of breast cancer or return a no recall decision (“all clear”) if not (Fig. 1). The comparison between human readers and the AI reader was performed at the screening episode level with the positive class defined as screen-detected cancers (biopsy-confirmed cancer at assessment within 6 months) and interval cancers (clients who develop breast cancer between six months after a screen and the date of their next screen), and the negative class defined as any client who does not develop cancer within the screening interval (12 or 24 months). We summarised reader and system performance on the test set with area under the receiver operating characteristic (ROC) curve (AUC), and sensitivity and specificity of cancer detection decisions at specific operating points.

**Figure 1.**
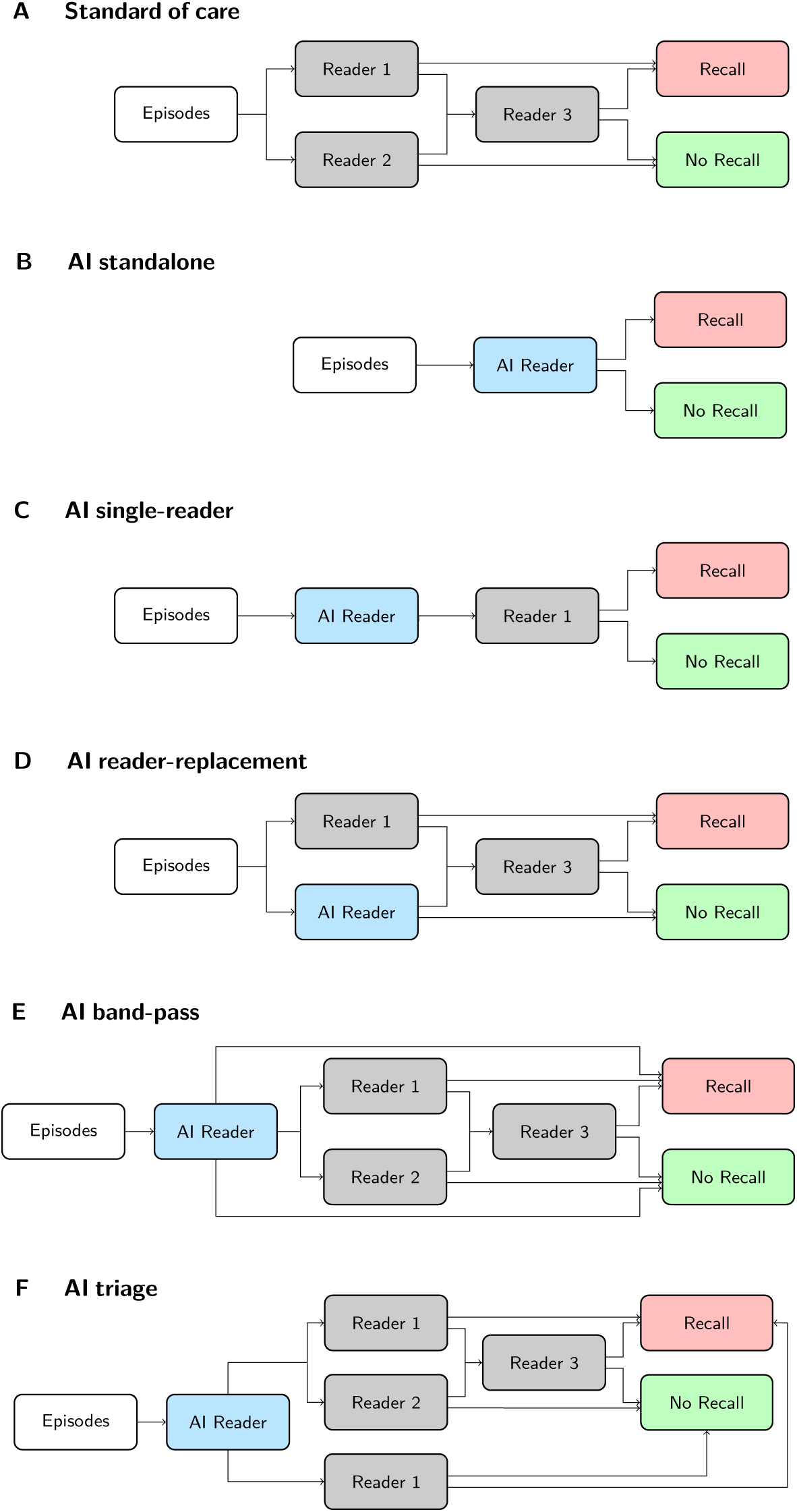
Screening episode flows for the current reader system and AI integration scenarios. (A) Standard of care scenario: Readers 1 and 2 see the same episode and opt to recall or not-recall, if they disagree Reader 3 arbitrates. (B) AI standalone scenario: all decisions are taken by the AI Reader without human intervention. (C) AI single-reader scenario: Reader 1 takes the final decision using AI Reader input. (D) AI reader-replacement: same as (A) but with AI Reader replacing Reader 2. (E) AI band-pass scenario: AI Reader screens out episodes before Readers 1 and 2. Episodes with high scores trigger the recall decision directly, and episodes with low scores trigger the no-recall decision directly. The other episodes continue to the usual reader system. (F) AI triage scenario: AI reader triages the episodes before Readers 1 and 2. Episodes with high scores continue to the usual system, and episodes with low scores go through the path with only 1 reader.

With detailed simulation studies, we compare the standard of care (two human readers with arbitration; Fig. 1A) to five AI-integrated screening pathways. We examine the AI reader as a standalone reader (AI standalone; Fig. 1B), as a reader aid for a single reader (AI single-reader; Fig. 1C), as a replacement for the second reader in a two reader with arbitration pathway (AI reader-replacement; Fig. 1D), as a filter for high confidence recall and no-recall decisions (AI band-pass; Fig. 1E), and as making a triage decision between a single reader and a two reader with arbitration pathway (AI triage, Fig. 1F). In the simulations our baseline reference point for system performance is the observed standard of care results from the original reads, with arbitration reads simulated from historical reader performance when needed. For each AI-integrated scenario we vary the AI reader operating points to identify settings for optimal performance. We then select candidate operating points for each scenario and vary the human reader performance by modelling an interaction effect of the AI reader on human readers. We simulate these interaction effects by changing human reader decisions to agree with the AI reader (varying the proportion between 0% and 50%) when the AI reader is correct (positive interaction), incorrect (negative interaction), and uniformly (neutral interaction, typically referred to as “automation bias”). More details on the datasets, screening scenarios, and the simulation design can be found in the Methods.

### AI as a standalone reader

We first evaluate the AI reader in the standalone scenario, assessing its performance when it replaces all human readers in the standard of care. The AI standalone scenario achieved an AUC of 0.932 (95% CI 0.923, 0.940) when evaluated on the retrospective test set (Fig. 2A). The AI standalone operating point achieved higher performance than the mean of individual radiologists (weighted by number of reads), in both sensitivity (75.0% vs. 66.3%; *P* < 4.11 × 10^−8^) and specificity (96.0% vs. 95.6%; *P* < 1.11 × 10^−9^). However, operating as a sole reader the AI reader had lower sensitivity (75.0% vs. 79.8%; *P* < 1.98 × 10^−5^) at non-inferior specificity (96.0% vs. 96.0%: *P* = 0.56) than the current standard of care (two readers with arbitration; Supplementary Table 3).

**Figure 2.**
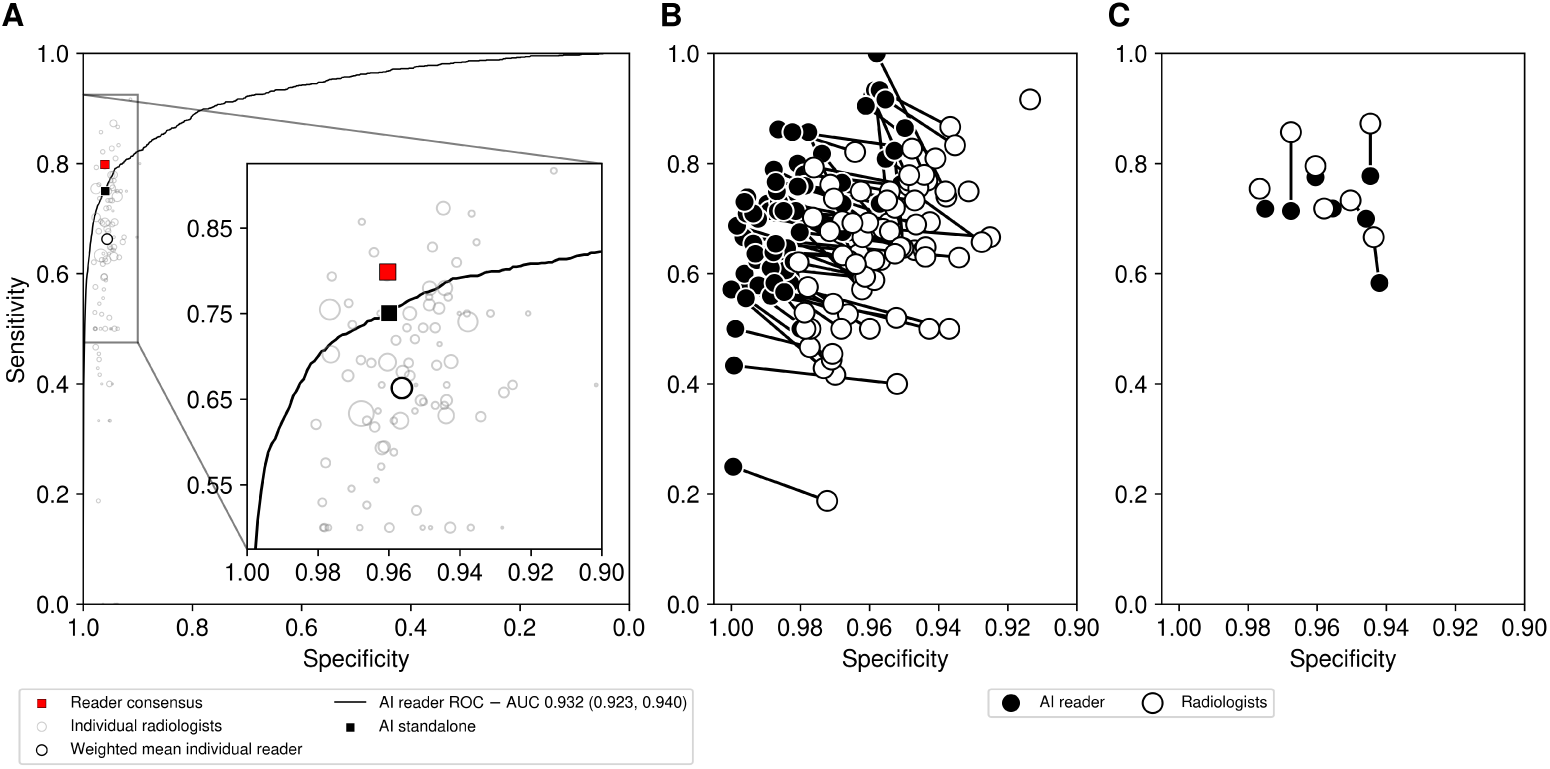
Performance of the AI reader on retrospective cohort. (A) The AI reader ROC curve compared with the weighted mean individual reader and reader consensus for 149,105 screening episodes. The AI reader achieved an AUC of 0.932 (95% CI 0.923, 0.940) above the weighted mean individual reader performance (95.6% specificity, 66.7% sensitivity) but below the reader consensus performance (96.1% specificity, 79.8% sensitivity; standard of care). The weighted mean individual reader (black circle) is the mean sensitivity and specificity of all the individual readers (grey circles; 125 readers) weighted by their respective total number of reads. (B) and (C) AI reader compared against 81 individual readers (min. 1,000 reads). An optimal point from each AI reader ROC curve is shown for each comparison. We show separately human readers for which both sensitivity and specificity of the AI reader point was greater than or equal to the reader (B; 74 readers, 91.3% of readers; 253,328 reads, 88.3% of reads) and readers for which the AI reader is less than or equal to the human reader in either sensitivity or specificity (C; 7 readers, 8.6%; 33,525 reads, 11.7%).

To better understand how the AI reader ranked amongst the reading cohort, we compared the AI reader against all individual readers in the first or second reader position with at least 1,000 reads and found that with well-chosen operating points the AI reader could achieve higher sensitivity and/or specificity than 74 out of 81 human readers (see Fig. 2B and C). When compared with the third readers on the third read cohort, the AI reader performance was found to be lower than that of the mean third reader (Supplementary Fig. 2). This result supports the use of the AI reader only in the first or second position, as we will explore in the other scenarios.

A key observation on AI reader performance was that the AI standalone scenario ROC curve was above that of the current standard of care for first round episodes (that is, the first time a client attended the screening service) and above the weighted mean reader for second and subsequent rounds (Supplementary Fig. 3). Further comparisons were performed across other breakdowns including age, manufacturer, and radiologist morphology labels (Supplementary Fig. 4; Supplementary Fig. 5). We also investigated the AI reader scores by outcome class label and found the top 10% of scores contained 92.7% of screen-detected cancers and 42.2% of interval cancers (81.8% of all cancers; Supplementary Table 1). Additionally, we benchmarked our AI reader on external datasets (some publicly available) achieving state-of-the-art performance (Supplementary Table 2).

Overall, the AI reader was a strong individual reader, outperforming individual readers on average. The first round screening results highlight the strength of the AI readers when limited additional information is available (specifically, prior screens or other reader opinions), while the third reader and second and subsequent round analysis shows how the human readers are able to make use of the additional information to improve performance. The AI standalone scenario did not outperform the standard of care but could offer improvement in single-reader settings.

### AI-integrated scenarios without human-AI interaction effects

Integrating the AI reader within a multi-reader pathway offers a more practical and clinically viable option than AI as a standalone reader, with multiple prospective trials underway globally [20, 23, 24]. We conduct detailed simulation studies of the AI reader-replacement, AI band-pass, and AI triage scenarios compared against the current standard of care in breast cancer screening in Australia, two independent readers with arbitration (Fig. 1; Methods). In this section, we vary only the AI reader operating points and simulate arbitration reads as required based on historical performance (Methods). For the AI band-pass and AI triage scenarios we assume the human readers are blinded to AI reader outputs and in the AI reader-replacement scenario the first reader is also assumed to be blinded.

The AI reader-replacement scenario produced higher system sensitivity (82.3%; 95% CI 81.5–83.1, *P* < 0.0025) and higher specificity (96.3%; 95% CI 96.2–96.3, *P* < 4.3 × 10^−6^) than the current standard of care system (Fig. 3A; Supplementary Table 4). Across 149,105 screening episodes in our retrospective testing set, the AI reader-replacement scenario had 354 fewer unnecessary recalls (−6%, false positives) and detected 33 more cancers (+3.1%, true positives) with a reduction of 147,959 human reads (−48%), but required 11.6% more third (arbitration) reads (Table 1; Supplementary Fig. 6). The modelled reading and assessment cost reduction was 15%.

**Figure 3.**
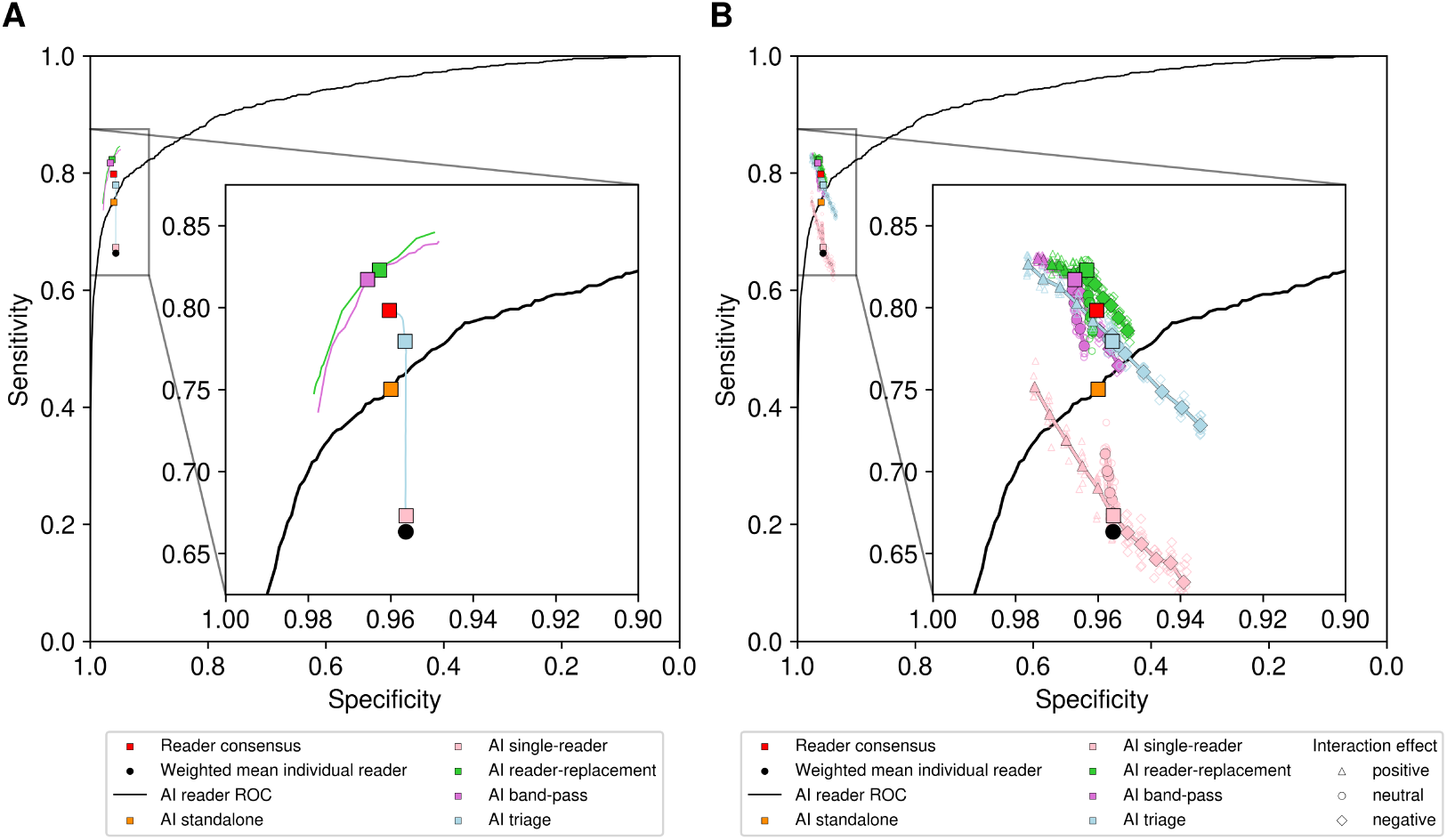
Comparison of AI integrated scenarios. (A) Human reader consensus performance compared with AI standalone, AI reader-replacement, AI band-pass, and AI triage on the retrospective cohort (149,105 screening episodes). Representative points are shown for AI standalone (96.0% specificity, 75.0% sensitivity), AI single reader (95.6% specificity, 67.3% sensitivity), AI reader-replacement (96.3% specificity, 82.3% sensitivity), AI band-pass (96.6%, 81.7%) and AI triage (95.7% specificity, 78.0% sensitivity). Other potential operating points are shown as a continuous line. Both AI reader-replacement and AI band-pass improved performance over the human reader consensus (96.1% specificity, 79.8% sensitivity). (B) AI integrated scenarios when reader performance is varied with an interaction effect when the human reader disagrees with the AI reader. From 0% to 50% of discordant decisions are reversed when the AI reader was correct (triangle, positive effect), uniformly (circle, neutral effect) and incorrect (diamond, negative effect). For AI triage to match human reader consensus performance a 15% positive interaction effect of the AI reader on human readers is required.

**Table 1.** Comparison of the current reader system and the AI-integrated scenarios by the screening outcomes and workload. For the AI reader replacement scenario, mean values based on 1000 simulations are presented (rounded to the nearest integer, and the 95% confidence intervals (CI) are presented for the sensitivity and specificity). The AI band-pass and AI triage scenarios do not require simulating the third reader, and the results are entirely based on the real data (i.e., no CI). Percentages in the brackets with the plus / minus sign indicate the percentage change compared with the current reader system.

| Variable | Current reader system | AI reader-replacement | AI band-pass | AI triage |
| --- | --- | --- | --- | --- |
| Sensitivity (%) | 79.8 | 82.3 (81.5, 83.1) | 81.7 | 77.2 |
| Specificity (%) | 96.0 | 96.3 (96.2, 96.3) | 96.6 | 95.7 |
| No. of episodes | 149105 | 149105 | 149105 | 149105 |
| True positive | 1061 | 1094 (+3.1%) | 1086 (+2.4%) | 1026 (-3.3%) |
| True negative | 141915 | 142269 (+0.2%) | 142694 (+0.5%) | 141490 (-0.3%) |
| False positive | 5861 | 5507 (-6%) | 5082 (-13.3%) | 6286 (+7.3%) |
| False negative | 268 | 235 (-12.4%) | 243 (-9.3%) | 303 (+13.1%) |
| Workload |  |  |  |  |
| Assessments | 6922 | 6602 (-4.6%) | 6168 (-10.9%) | 7312 (+5.6%) |
| Human reads | 308091 | 160132 (-48%) | 59453 (-80.7%) | 166292 (-46%) |
| *Third reads | 9881 | 11027 (+11.6%) | 3173 (-67.9%) | 2276 (-77%) |

\*The third reads are part of the human reads. The separate entry is presented to show the workload impact on the third reader.

The AI band-pass screening scenario also achieved both higher system sensitivity (81.7%, *P* < 0.0058) and specificity (96.6%, *P* < 2.2 × 10^−25^) than the current standard of care system (Fig. 3A). The AI band-pass screening scenario had 779 fewer unnecessary recalls (−13.3%) and detected 25 more cancers (+2.4%) with a reduction of 248,638 human reads (−80.7%), while also providing a 67.9% reduction in third reads (Table 1; Supplementary Fig. 7). The modelled reading and assessment cost reduction was 28.3%.

The AI triage screening scenario, without modelled AI-human interaction, produced lower sensitivity (77.2%) and specificity (95.7%) than the current reader system (Fig. 3A). Using an operating point to shift 90% of reads to a single-reader pathway, the AI triage screening scenario had 425 more unnecessary recalls (+7.3%) and detected 35 fewer cancers (−3.3%) with a reduction of 141,799 human reads (−46%), including a 77.0% reduction in third reads (Table 1; Supplementary Fig. 7). The modelled reading and assessment cost reduction was 7.5%. This result is expected as AI triage allocates a proportion of reads to a single-reader pathway, which will always result in reduced performance in the absence of human-AI interaction.

Both the AI reader-replacement and AI band-pass scenarios offer opportunities for improved performance relative to the current standard of care across different operating points, with AI reader-replacement achieving the highest reduction in missed cancers (−12.4%) and the AI band-pass having the highest reduction in unnecessary recalls (−13.3%). The AI triage scenario offers good performance with a large workload reduction while maintaining a human decision maker for all episodes (which AI band-pass lacks). These retrospective simulations do not take into account the change in cancer prevalence [25] that both AI band-pass and AI triage would have on the episodes diverted to different pathways nor do they consider the potential positive and negative impacts of human readers having access to AI reader outputs. Human-AI interaction effects are explored in the next section.

### AI-integrated scenarios with human-AI interaction

We present three types of human-AI interaction by considering neutral, positive, and negative interactions in the AI integrated scenarios. In the AI reader-replacement scenario the interaction affects the arbitration reader only. Conversely, in the AI band-pass and AI triage scenarios, the interaction affects all readers. In the AI single-reader scenario, human-AI interaction affects the single reader operating with decision support.

In all cases, we are modelling an effect of human readers revising their decision when in disagreement with the AI reader. We vary the degree of how often the human reader revises their decision upon disagreement, ranging from 0 to 1, where 0 refers to the human reader never revising their decision and 1 refers to the human reader always revising their decision to agree with the AI (and 0.5 would refer to the human reader revising the decision 50% of the time). In the special case where the human reader changes their decisions to agree with AI fully, the collaboration between human reader and AI reader would collapse to the AI standalone scenario. For all scenarios, we use the AI standalone operating point when considering disagreement.

The neutral human-AI interaction case, in which a reader will be inclined to agree with the AI model irrespective of the correctness of its outcome, models a general form of “automation bias”. Automation bias is the tendency of humans to overly agree or rely on an automated system (the AI reader in our case) [26]. The positive human-AI interaction case refers to the human reader changing their decision to agree with the AI reader only when the AI reader is correct, simulating human readers accurately discriminating between useful and spurious AI reader outputs. The negative case refers to the human reader changing their decision only when the AI reader is incorrect.

### AI reader-replacement

In the AI reader-replacement scenario, assuming positive human-AI interaction, AI improves the system’s specificity but has minimal impact on sensitivity (the green triangles in Fig. 3B). This is because the AI reader operates with higher specificity (96.0%) and lower sensitivity (75.0%) compared to the third reader (56.3%, 97.5%). At this operating point, AI only correctly identifies a few cases of cancer missed by human readers, and human readers do not benefit much from it. Conversely, in the negative scenario, as Reader 3 is highly sensitive and AI is highly specific, agreeing with AI leads to a substantial loss of sensitivity but not specificity. With negative interaction, Reader 3 is more prone to overlooking cancer cases rather than misidentifying normal cases when learning from the AI reader.

In the neutral case, agreeing with the AI would gradually reduce the multi-reader system to a one-reader system, where the decision is solely driven by the AI reader. As a result, the performance would tend towards the AI standalone scenario at (75.0%, 96.0%). Nevertheless, in the AI reader-replacement scenario roughly 30% or more of discordant decisions would need to be reversed in the neutral and negative interaction cases for system sensitivity to drop below that of the current standard of care (reader consensus). In other words, the AI reader-replacement scenario, without interaction effects, is so effective that human readers could accept up to 30% of the AI’s mistakes before its performance falls below the standard of care, making it substantially robust to the downside risks of human-AI interaction.

### AI band-pass

For the band-pass scenario with positive human-AI interaction effect, AI improves on specificity and sensitivity, more so than in the AI reader-replacement scenario because all three readers may benefit from AI outputs. In the negative case, the opposite is true if human reader performance suffers due to the mistakes made by AI. The sensitivity decreases significantly, with most of the contribution coming from Reader 3. The specificity also decreases slightly but remains at approximately 96%. In general, the AI reader tends to return more false negatives than false positives due to its high specificity.

In the neutral case, agreeing with AI would gradually lead to the collapse of the mid-band two-reader-and-consensus system into the AI standalone scenario. As the interaction effect increases, the sensitivity and specificity of the band-pass scenario will approach that of the AI standalone scenario. But unlike the AI reader-replacement scenario, the transition may not be complete, as the high-band and low-band paths continue to function, even if they only contain a limited number of episodes.

### AI triage

The AI triage scenario shows the widest range in performance of the multi-reader scenarios, depending on the nature of the human-AI interaction. When there is a positive interaction, AI triage would achieve the baseline consensus (standard of care) performance when humans benefit from the AI’s ability to correct 15% of the decisions in cases of disagreement. The potential benefits are substantial: if all human readers adopt 50% of the AI’s corrections, the sensitivity could increase by 5.3 percentage points compared to the baseline where there is no interaction between humans and AI. This would result in a performance that exceeds the baseline standard of care in terms of sensitivity by 3.4 percentage points, while also operating at 97.7% specificity. In the negative interaction case, however, the potential downside is significant. The sensitivity can quickly drop by 5.6 percentage points to about 73.1% (6.7 percentage points lower than baseline standard of care) and the specificity can drop to 93.5% when human readers follow the mistakes made by the AI reader. In the neutral case, agreeing with the AI reader does not change the performance significantly, and so performance remains a little below standard of care in both sensitivity and sensitivity.

The performance of the AI triage scenario presents large variations when subject to positive or negative interactions, because human readers are responsible for all the recall decisions, while the AI merely supports them. When the AI offers beneficial assistance, the entire scenario improves as all readers gain from it. Conversely, if the assistance is negative, the scenario worsens in a similar way. In the neutral case, the scenario does not improve because we know from the AI standalone case that the AI reader is tuned to match the reader at about the same specificity but with much improved sensitivity. However, the one-reader pathway contains all the low-scoring and mostly normal episodes, so improved sensitivity does not have an effect. And since the three-reader pathway has high sensitivity, the AI assistance does not detect more cancers. In this case, triage is not as susceptible to automation bias, as agreeing with AI would not harm the system’s performance.

The AI triage scenario provides a distinct contrast to the AI reader-replacement scenario. The positive and negative interactions have less impact in the AI reader-replacement scenario than in the triage scenario, because the interaction applies to fewer human readers. Also, the AI reader serves two roles in the reader-replacement scenario: as a screening tool, where it improves the system beyond the consensus performance, and as a diagnosis assistive tool. Since it has already delivered most benefits as a screening tool, the additional improvements from its role as an assistive tool are relatively minor. The triage scenario, on the other hand, receives the impact of human-AI interaction for all human readers and the structure of the pathway means that the AI has strong effects both as a screening tool and as an assistive tool.

### AI single-reader

In the AI single-reader scenario, with positive interaction correcting up to 50% of the original decisions, the system can achieve a sensitivity of 74.6%, on par with the AI standalone, but with an additional 1.6% improvement in specificity (Fig. 3B). For the negative interaction case, with up to 50% of the discordant decisions turning into errors, the sensitivity and specificity decrease to approximately 62.6% and 93.9% respectively. There are more gains than losses in this scenario, because disagreements occur more frequently when AI is correct than when it is wrong. In the neutral case, as the human reader increasingly agrees with the AI reader, the performance tends towards the AI standalone scenario. The automation bias turns out to be favourable here, because the AI reader outperforms the average reader by a sizable margin, and in a one-reader pathway this directly translates to an improved system performance. Across all scenarios, a single-reader AI system is unable to match the performance of a multi-reader system when it comes to screening outcomes. However, using AI significantly narrows the gap between the single-reader system and the two-reader-and-consensus system, making a compelling case for consideration in settings limited to single-reader screening pathways.

## Discussion

In this study we used detailed simulations to evaluate how an AI reader for breast cancer detection could perform in different single- and multi-reader settings in population mammographic screening. We explored positive, neutral, and negative human-AI interaction effects and identified the major upside and downside possibilities for four AI-integration scenarios depending on the nature and strength of the interaction effects. The AI reader used was a strong individual reader trained on the ADMANI training datasets [27], with its performance assessed on a carefully selected and unseen ADMANI testing dataset. This AI reader achieved significantly higher sensitivity (+8.3%) and specificity than the weighted mean individual human reader, and better performance than 91% of individual human readers in our retrospective testing dataset. ă

The AI standalone reader’s high performance and minimal running cost (it is fully automated so eliminates costs associated with human readers) make a compelling case for its use in settings that follow single-reader screening practices. Many countries currently implement single-reader screening, whether for reasons of historical choices to prioritise cost and operational efficieny, as in the United States, or because the resources have not been available to establish a multi-reader screening pathway integrated into healthcare systems, as in many other countries. In such settings, AI readers could play an important role in improving screening system performance while minimising costs. However, despite the AI reader’s impressive performance as a single reader, as a standalone system the AI reader cannot match the performance of the current standard of care (two readers with arbitration) used in Australia and many other countries including Sweden and the UK.

This performance gap necessitates some human-AI collaboration to improve screening outcomes [28] and, furthermore, practical, social, and legal considerations encourage retaining the human central to the decision making process [22]. We observe that the AI reader can outperform human readers in the first/second position, but not in the third reader role. This observation suggests that there is a preferred position for the AI reader to maximise its advantages relative to human readers: it should serve in a more “junior” role where its excellent specificity optimises the performance of the whole screening pathway. Relatedly, the AI reader works extremely well for first-round screening, outperforming the standard of care in this study dataset.

We studied four collaborative AI-integrated scenarios for the screening pathway: AI single-reader, AI reader-replacement, AI band-pass, and AI triage. Without any human-AI interaction effects, both AI reader-replacement and AI band-pass demonstrated significantly superior sensitivity and specificity compared to the standard-of-care two reader with arbitration system. Interestingly, these two approaches achieve this superior performance with different characteristics. The AI reader-replacement system gains most from reducing missed cancers (false negatives), while the AI band-pass system gains most from limiting unnecessary recalls to assessment (false positives). Both the AI single-reader and AI triage systems demonstrate lower performance than the standard of care and other multi-reader AI-integrated systems. However, if we assume that positive human-AI interaction yields a 15% improvement in human reader decision making, then AI triage can match the current standard of care. Furthermore, the AI triage system has the highest possible upside if there are strong positive human-AI interaction effects. Interim results from the ongoing MASAI trial provide evidence that positive human-AI interaction is plausible [20]. For the AI single-reader, the superior performance of the AI reader over human readers provides a safety net, leading to improved performance even with neutral reliance on the AI reader. The relative gains from the use of AI are greater in the single-reader pathway than in the multi-reader pathways thanks to the strong performance of the AI reader as a single reader relative to single human readers. The gap between the single-reader pathway and multi-reader pathways is almost halved when AI is used in both.

The four AI-integrated scenarios present differing considerations when it comes to implementation and clinical application. AI reader-replacement is conceptually straight-forward and is the least disruptive to the reader system as it retains a human decision maker for all episodes. As the overall reader structure is unchanged, every episode must have at least one human reader, the cancer prevalence for the first reader remains the same, and only the third reader can view AI outputs. The limited exposure of readers to the AI reader outputs mitigates the risks of negative human-AI interaction, but also limits the upside of positive human-AI interaction. The AI band-pass scenario offers perhaps the greatest potential for tuning the pathway, allowing for improved performance while minimising radiologist workload. However, having an AI reader make the final reading decision, without human involvement, faces challenges before clinical adoption. Concerns relating to bias, quality assurance, and medico-legal responsibility would need to be addressed [22]. With strong performance, the AI reader-replacement and AI triage systems seem most promising for clinical implementation as their “human-in-the-loop, human-in-charge” architectures minimise barriers to their adoption.

Our study features limitations based on constraints on what is feasible in complex simulations using a retrospective cohort, and other factors and considerations beyond the scope of this study. Here, we did not account for potential effects of a change in cancer prevalence that readers in both AI triage and AI band-pass will experience. We also consider positive, neutral, and negative human-AI interaction effects in isolation, and only for a single AI reader operating point. Human readers are also likely to modulate their decisions based on scores or other supporting information, which we did not consider. Alongside prospective studies and further development of the datasets, translation into screening programs will require, or be aided by, other programs of work not covered in this study, including: (1) Development of approaches to algorithm quality assurance to assess bias and drift on an ongoing basis; (2) Deeper examination of AI explainability for client and clinician, and their acceptance of AI reader false negatives and positives; (3) Further development of algorithms to focus on interval cancers (false negatives); (4) Utilising AI to predict short-, medium- and long-term breast cancer risk and to support personalized screening pathways. AI reader performance may be improved by integrating a longitudinal view of episodes for a given client over multiple screening rounds, taking advantage of information from prior screens as human readers do.

Taken together, we have shown, on a large retrospective dataset under different scenarios and conditions, how an AI reader can be integrated into breast cancer screening programs to improve cancer detection, minimise unnecessary recall to assessment, and lower human reader workload and cost. Our work extends previous research by simulating arbitration reads and human-AI interaction, as well as conducting thorough analysis and comparison across various AI integrated screening pathways using a common large retrospective dataset and AI system. Our results provide insights on optimising breast cancer screening outcomes through AI positioning and pathway design. Overall, our simulation results provide evidence that supports the prospective evaluation of the AI integration pathways studied here and offers plausible approaches to clinical implementation of AI readers in breast cancer screening in the near future.

## Contributors

Conceptualization: HMLF, DJM, JL, JH, GC; Methodology: HMLF, CAPS, CFK, ME, YC, CW, PB, GC, DJM; AI model and code development: CAPS, CFK, ME, YC, CW; Dataset development: HMLF, ME, CAPS, CFK; Data analysis: CAPS, CFK, ME; Resources: DJM, HMLF, PB, JL, JH, GC; Writing - original draft, review & editing: CAPS, CFK, ME, HMLF, DJM, GC; Supervision: HMLF, DJM, GC; Project administration: HMLF, DJM, GC; Funding acquisition: HMLF, DJM, PB, JL, JH, GC.

The BRAIx team comprises: Helen ML Frazer^1,2^, Carlos Andres Peña-Solorzano^3,4^, Chun Fung Kwok^3,4^, Michael S Elliott^3,4^, Yuanhong Chen^5^, Chong Wang^5^, Prabhathi Basnayake Ralalage^1^, Ravishankar Karthik^2^, Katrina Kunicki^3^, Daniel Schmidt^7^, Enes Makalic^7^, Osamah Al-Qershi^7^, Prue C Weideman^7^, Samantha K Fox^7^, Shuai Li^7^, Tuong L Nguyen^7^, Brendan Hill, Jocelyn F Lippey^1,6^, John L Hopper^7^, Peter Brotchie^8^, Gustavo Carneiro^5^, Davis J McCarthy^3,4^. ^1^St Vincents BreastScreen, St Vincents Hospital Melbourne, Victoria, Australia; ^2^BreastScreen Victoria, Victoria, Australia; ^3^Bioinformatics and Cellular Genomics Unit, St Vincent’s Institute of Medical Research, Victoria, Australia; ^4^Melbourne Integrative Genomics, School of Mathematics and Statistics/School of BioSciences, Faculty of Science, University of Melbourne, Victoria, Australia; ^5^School of Computer Science, Australian Institute for Machine Learning, University of Adelaide, South Australia, Australia; ^6^Department of Surgery, St Vincents Hospital Melbourne, Victoria, Australia; ^7^Centre for Epidemiology & Biostatistics, Melbourne School of Population and Global Health, University of Melbourne, Victoria, Australia; ^8^Department of Radiology, St Vincents Hospital Melbourne, Victoria, Australia.

## Declaration of interests

Peter Brotchie is an employee of annalise.ai.

## Data availability

The non-transformed image and non-image data that established the ADMANI datasets were accessed under license agreement with BreastScreen Victoria. Further details about the ADMANI datasets are available in the data descriptor paper [27]. The three datasets used as external validation are publicly available or available via request. The Chinese Mammography Dataset (CMMD) is publicly available from the following website: https://wiki.cancerimagingarchive.net/pages/viewpage.action?pageId=70230508. The Cohort of Screen-age Women - Case control (CSAW-CC) dataset is available via request from the following website: https://snd.gu.se/en/catalogue/study/2021-204. The Breast Screen Reader Assessment Strategy Australia (BREAST Australia) is available via request from the following website: https://breast-australia.sydney.edu.au/research/.

## Code availability

The code used for training the BRAIx AI reader is based on open-source algorithms and training techniques. We are required to protect potentially commercially valuable project intellectual property, which the source code constitutes, as part of our multi-institution agreement and grant obligations. The description of the model training procedure and models used are provided in the Methods section and can be implemented with open-source frameworks. The main conclusions drawn in our work relate to our AI reader simulation experiments. The code used to simulate the AI reader operating within the screening program and validate the external results are available publicly at https://github.com/BRAIx-project/retrospective-cohort-study.

## Acknowledgments

The authors would like to acknowledge Rita Butera, Luke Neill, and Georgina Marr from BreastScreen Victoria and the leadership and staff of St Vincent’s Hospital Melbourne for their support of the project. The authors would like to acknowledge Katrina Kunicki, Anne Johnston, Colleen Elso, Liz Campbell and Tom Kay of St Vincents Institute of Medical Research (SVI) for their extensive help and support with many aspects of this project. The authors thank Wayne Crismani for thoughtful comments on the manuscript. SVI provided IT support, infrastructure, and GPUs used for numerical calculations in this paper enabled via funding from the Victorian State Government Operational Infrastructure Support Program to St Vincents Institute of Medical Research. HMLF acknowledges the generous support of the Royal Australian and New Zealand College of Radiologists (Clinical Research Grant), St Vincent’s Hospital Melbourne (Research Endowment Fund) and The Medical Device Partnering Program for enabling the prior foundational research to be undertaken. DJM acknowledges generous support from Paul Holyoake and Marg Downey.

## Methods

### Inclusion and ethics

The conceptualisation, design, and implementation of this study was conducted with close collaboration between clinical staff working in the organised population breast screening service in Victoria, Australia and local academic researchers. Ethics approval for this study was provided by the St Vincent’s Hospital, Melbourne human research ethics committee (approval no. LNR/18/SVHM/162). All women sign a consent form at screening registration that provides for the use of the de-identified data for research purposes. A unique identifier is used for the purposes of the ADMANI datasets, with all image and non-image data de-identified.

### Screening program

The BreastScreen Victoria screening program is a population screening program open to women aged 40+ with those between the ages 50–74 actively recruited. A typical BreastScreen Victoria client has a mammogram taken with a minimum of four standard mammographic views (left and right mediolateral oblique, MLO, and craniocaudal, CC) every two years. Annual screening is offered to a small proportion of high-risk clients (<2%).

Every client undergoing screening through BreastScreen Victoria experiences a standardised screening pathway and data generation process (Supplementary Fig. 8). Each mammogram is read independently by two breast imaging radiologists who indicate suspicion of cancer, all clear, or technical rescreen. If there is disagreement a third reader, with visibility of the original two readers’ decisions, determines the final reading outcome. Clients with a suspicion of cancer are recalled for assessment. At assessment, further clinical workup and imaging is performed. Any client who has a biopsy-confirmed cancer at assessment (within six months of screening) is classified as a screen-detected cancer (true positive). Any clients who are recalled but confirmed with no cancer after follow-up assessments are classified as either benign or no significant abnormality (false positive). Clients who were not recalled at reading and do not develop breast cancer within the next screening interval are classified as normal (true negative). Clients who develop breast cancer between six months after a screen and the date of their next screen (12 or 24 months) are classified as interval cancers (false negative). The datasets we use are structured around individual screening episodes of clients attending BreastScreen Victoria. A screening episode is defined as a single screening round that includes mammography, reading, assessment, and the subsequent screening interval.

### Study Datasets

The datasets used in this study were derived from the ADMANI datasets [27]. The ADMANI datasets comprise of 2D screening mammograms with associated clinical data collected from 46 permanent screening clinics and two mobile services across the state of Victoria, Australia. The entire datasets span 2013-2019, 2013-2015 were cancer enriched samples and not used for testing, 2016-2019 were complete screening years containing all episodes. Screening episodes that were missing any of the standard mammographic views (left and right MLO and CC), had incomplete image or clinical data, were were excluded (Fig. 4). If a screening episode had multiple screening attempts only the final attempt was used. Clients with breast implants or other medical devices were included. After exclusions a random 20% sample of all screening clients who attended between 2016-2019 were used in the study, the remaining 80% of clients were used in model training and development. The study dataset was further split into testing (75% of clients in study) and a development dataset (25% of clients in study) on which operating points were set. The testing dataset comprised 149,105 screening episodes from 92,839 clients and the development dataset 49,796 screening episodes from 30,946 clients (Table 2).

**Figure 4.**
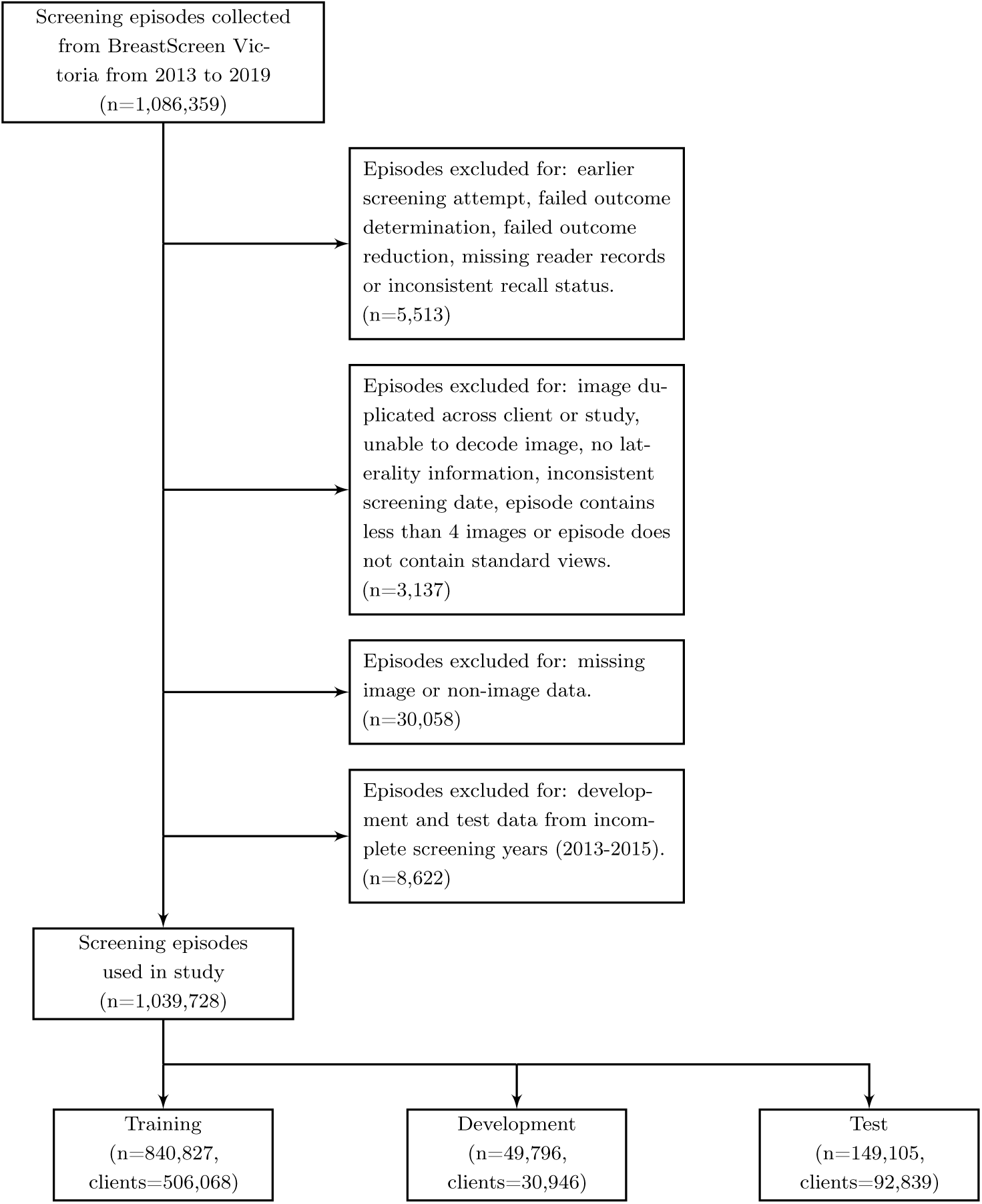
Screening episode exclusion criteria. Flow diagram of study exclusion criteria for screening episodes from the standardised screening pathway at BreastScreen Victoria. Missing data could be clinical data without mammograms or mammograms without clinical data, clinical data could also be incomplete missing assessment, reader or screening records. ‘Earlier screening attempt’ refers to a client returning for imaging as part of the same screening round, only the last attempt was used. ‘Failed outcome determination’ and ‘failed outcome reduction’ refer to being unable to confirm final screening outcome for the episode. ‘Missing reader records’ refers to missing reader data. ‘inconsistent recall status’ refers to conflicting data sources on whether episodes was recalled. ‘Incomplete screening years’ refers to years in which we did not have the full year of data to sample from (2013-2015), these years were excluded from testing and development datasets as they are not representative.

**Table 2.**
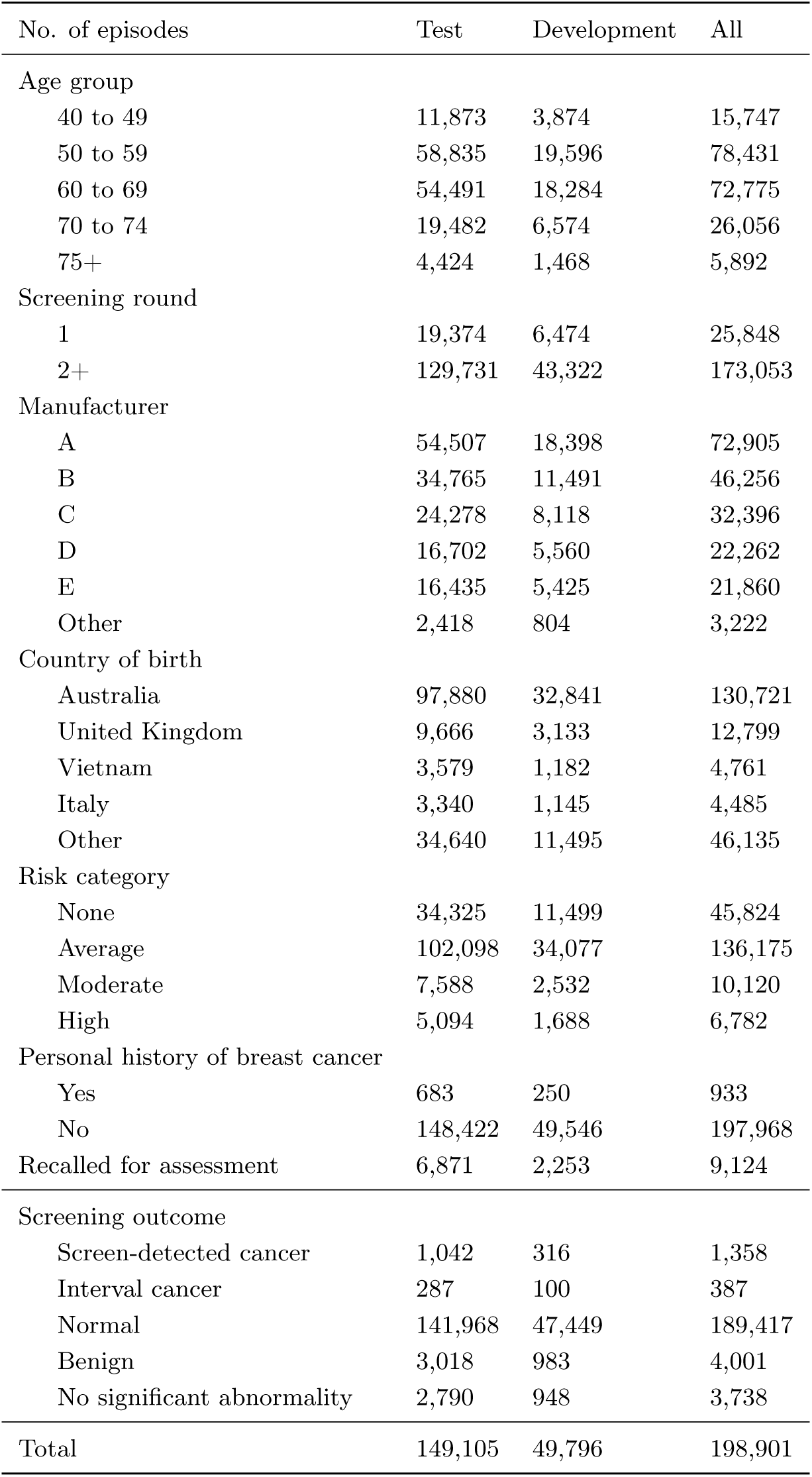
Summary and characteristics of data used in the study.

The study dataset has strong ground truths for all cancer (screen detected and interval) and non-cancer (normal, benign, or no significant abnormality (NSA) with no interval cancer). Cancer was confirmed by histopathology for screen-detected cancers or obtained from cancer registries for interval cancers. The histopathological proof was predominantly from an assessment biopsy confirmed with subsequent surgery. The ground truth for clients without cancer was a non-cancer outcome after reading and no interval cancer (normal) or non-cancer outcome after assessment and no interval cancer (benign or NSA). Information on country of birth, whether or not the client identifies as Aboriginal and/or Torres Strait Islander, and age was collected at the time of screening. Responses for country of birth and Aboriginal and/or Torres Strait Islander identification were aggregated into categories of First Nations Australians and regions.

Separately to the retrospective analysis a prospective dataset was collected from December 2021 to May 2022. Data was collected in real-time (daily) from a single reading and assessment unit (St Vincent’s Breastscreen, Melbourne, Australia) using two mammography machine manufacturers from December 2021 to May 2022. The prospective dataset contains the same ground truth and demographic information with the exception of interval cancer data as it was not yet available at the time of publication. The prospective dataset consisted of a total of 25,848 episodes and 108,654 images from 25,848 clients with a total of 195 screen-detected cancers (Supplementary Table 5).

### AI Reader System

For this study we used the BRAIx AI Reader (v3.0.7), a mammography classification model developed by the BRAIx research program. The model is based on an ensemble of modern deep learning neural networks and trained on millions of screening mammograms. We studied and created an ensemble from ResNet [29], DenseNet [30], ECA-Net [31], EfficientNet [32], Inception [33], Xception [34], ConvNext [35], and four model architectures developed specifically for our problem, including two multi-view models that use two mammographic views of the same breast concurrently [36], and two single-image interpretable models that provide improved prediction localisation and interpretability [37]. Each model from the ensemble was implemented in PyTorch [38] and trained on data splits from the training set. The models were trained for 10-20 epochs using the Adam optimiser [39] with an initial learning rate of 10^−5^, with weight decay of 10^−6^, and with the AMSGrad variant enabled [40]. The training set was selected to have about 10:1 ratio for non-cancers (benign, no significant abnormality and normal) and screen-detected cancers, respectively. To enforce a specific ratio, not necessarily all the available non-cancer images in the dataset are used during training of the models. Images were pre-processed to remove text and background annotations outside the breast region, then cropped and padded to keep the same height-to-width ratio of 2:1. Data augmentation consisted of random affine transformations [41].

The AI reader is image-based and produces a score associated with the probability of malignancy for each image. Image scores are combined to produce a score for each breast and the maximum breast score is the episode score. Decision thresholds convert each episode (or breast) scores to a recall or no-recall decision. There are no minimum number of images required. Elliptical region-of-interest annotations are produced from the pixels that contribute most to the classification score and multiple regions are ranked by importance (Supplementary Fig. 9). The reader has been evaluated on publicly available international datasets and achieved state-of-the-art performance (Supplementary Table 2). The distribution of episode scores from the study dataset, useful for inter study comparisons, are also available (Supplementary Table 1).

### Simulation design, operating points and evaluation metrics

To provide insights into the AI reader and its potential in clinical application, we performed retrospective simulation studies, where we evaluated the AI reader performance as a standalone reader and in three AI-integrated screening scenarios.

### AI-integrated screening scenarios

Five scenarios were considered to evaluate the AI reader integrated in the screening pathway, AI standalone, AI single-reader, AI reader-replacement, AI band-pass, and AI triage. In the one-reader pathway, the reader makes a decision on all episodes, and the decision is final. This includes the AI standalone and the AI single-reader scenarios. In the two-reader-and-consensus pathway the first two readers individually make a decision on whether or not to recall the client for further assessment. If the two readers agree, that is the final reading outcome. If they disagree, a third reader, who has access to the first two readers’ decisions and image annotations, arbitrates the decision. This pathway includes AI reader-replacement, AI band-pass and AI triage scenarios.

In the AI standalone scenario, the AI reader replaces the (only) human reader in the one-reader pathway and provides the same binary recall or no-recall outcome as the human readers on all episodes. In the AI single-reader scenario, the AI reader acts as an assistive tool to human readers. It provides the binary recall or no-recall outcome to the human reader, but it does not make any decision on its own. The human reader with access to the AI output would first make a decision and then consider whether to revise its decision should there be disagreement with the AI output.

In the AI reader-replacement scenario, the AI reader replaces one of the first two readers in the screening pathway and provides the same binary recall or no-recall outcome as the human readers. The first and second readers are replaced at random (with equal probability) for each episode. As the AI reader could trigger a third read that did not exist in the original dataset, the third reader was simulated for all episodes even if an original third read was present. This approach is to prevent unduly tying the result to the dataset and to obtain better variability estimates. Sensitivity analysis where the third reader uses the real data when possible and where the replaced second reader is used as the third reader was also performed (Supplementary Table 6). The third reader in our retrospective cohort operated with a sensitivity of 97.5% and a specificity of 56.3% and we simulated the third reader with respect to this performance. Concretely, whenever an episode reaches the simulated third reader, the reader will make a recall decision by first inspecting the actual episode outcome and then using it as prediction 97.5% of the time if the outcome is cancer (and 2.5% of the time using the opposite case as prediction), or 56.3% if the outcome is normal. This is achieved by sampling from the uniform(0,1) distribution with the corresponding probability, and it ensures that the simulated performance matches the real-world performance. Confidence intervals were generated through 1000 repetitions of each simulation.

As a remark, we emphasise that the simulation of the third reader should be performed with reference to the real-world Reader 3, rather than by reusing data of the earlier replaced readers (e.g. Reader 1 or Reader 2) [19, 21]. Reusing data is convenient, as the replaced readers have seen all the episodes and it avoids the simulation of the third reader. However, this overlooks the fact that while Reader 1 and 2 make independent judgements, they are conditionally dependent. In simple terms, a difficult cancer case is difficult for any reader. In such cases, the two readers would frequently miss together even when they make independent judgement, and the overall sensitivity would drop if either of them is used as an arbiter. In general, Reader 3 (the arbiter) makes decisions differently than Readers 1 and 2 because Reader 3 has access to their decisions and analyses. If Readers 1 and 2 were used in place of Reader 3 for simulation, then the result would be distorted, as we see in Supplementary Table 6.

In the AI band-pass scenario, the AI reader was used analogously to a band-pass filter. The AI reader provided one of three outcomes: recall, pass, and no-recall. All episodes with recall outcome were automatically recalled. All episodes with the no-recall outcome were not recalled. All episodes with the pass outcome were sent to the usual human screening pathway. The AI reader made the final decision on the recall and no-recall episodes with no human reader involvement, and for all episodes that passed to the human screening pathway the original reader decisions were used.

In the AI triage scenario, the AI reader triages the episodes before the human readers. Episodes with high scores continue to the standard pathway, and episodes with low scores go through the pathway with only 1 reader. For episodes sent to the standard pathway, the original reader decisions were used, and for episodes sent to the single-reader path, the reader decision is sampled randomly (with equal probability) from the first and second readers. The AI reader made no final decision on any of the episodes.

### AI operating points

Three sets of operating points were used as part of the study: the AI reader-replacement reader, the AI band-pass reader and the AI triage reader. There are three sets for five scenarios because the AI standalone reader and the AI single reader uses the same operating point as the AI reader-replacement reader. All operating points were set on the development set (Supplementary Fig. 1).

The AI reader-replacement operating point used a set of manufacturer-specific thresholds to convert the prediction scores into a binary outcome: recall or no-recall. The operating point was chosen to improve on the weighted mean individual reader’s sensitivity and specificity. The weighted mean individual reader was the weighted (by number of reads) mean of the sensitivity and specificity of the individual (first and second) radiologists when they were operating as a first or second reader. For all operating points that improved on the weighted mean individual readers sensitivity and specificity the point with the maximum Youden’s index [42] was chosen.

The AI band-pass reader used two sets of manufacturer-specific thresholds to convert the prediction scores into three outcomes: recall, pass and no-recall. The AI band-pass simulation was evaluated at different AI reader thresholds via a grid search. At each evaluation, two thresholds for each manufacturer were set to a target high specificity and high sensitivity point. All episodes with a score above the high sensitivity point were given the recall outcome, all episodes below the high specificity point the no-recall outcome, and all episodes in between points were given the pass outcome. The final AI band-pass reader thresholds were chosen from the simulation result with the maximum Youden’s index of the points with non-inferior sensitivity and specificity than that of the two reader with arbitration system.

The AI triage reader used the 90% quantile of the prediction scores as the threshold to convert the prediction scores into the triage outcome: the standard pathway or the one-reader pathway. Episodes with prediction scores less than the threshold are assigned to the one-reader pathway; otherwise, they are assigned to the standard pathway.

When there is interaction between the AI reader and the human reader, i.e., human may revise their decision based on the AI output, the AI reader in all cases uses the reader-replacement operating point (which is also the standalone operating point). To clarify, taking AI triage scenario as an example, the AI triage reader uses the triage operating point to decide whether a episode should go to the standard pathway or the one-reader pathway. Once that is decided and the episode reaches any reader, the reader would have access to an AI assist reading tool that operates at the reader-replacement operating point. So overall, there would be two operating points functioning at the same time.

### Human-AI interaction

We simulate three interaction effects, the positive, the neutral and the negative effect. All three interactions involve an AI reader and a human reader. The AI reader first makes a decision about recall (using the assistive operating point), and then the human reader makes their decision with access to the AI output. The human may adjust their decisions if they differ from the AI’s, and this happens (100 × *p*)% of the time, where *p* is a parameter that varies between 0 and 1 across multiple simulations. For example, when *p* = 0.1, human readers will adjust the decision 10% of the time when their decisions differ from the AI, and when *p* = 1, human readers will change all their decision to align with the AI. This models the automation effect, which we refer to as the neutral interaction.

For positive interactions, the human readers would only adjust the decision if the AI is correct. This models the situation where AI enhances human readings by reducing occasional misses and assisting in complex cases. And for negative interactions, the human readers would change the decision only if the AI is incorrect. This models the situation where human is confused by the AI reader’s output and mistakenly changes their correct decision into an incorrect one.

### Evaluation metrics

The AUC, based on the receiver operating characteristic or ROC curve, is used to summarise the AI reader’s standalone performance.

Sensitivity and specificity are used to compare the AI reader with the radiologists and the AI-integrated screening scenarios with the current screening pathway. Sensitivity, or the true positive rate (TPR), is computed by dividing the number of correctly identified cancers (the “true positives”) by the total number of the observed cancers (“all positives”, i.e., including both screen-detected cancers and interval cancers). It measures the success rate of the classifier in detecting the cancer. This is a key performance metric because early detection of cancer leads to more effective treatment [43], due to a timely intervention, requirement for a less aggressive treatment, and improved survival rates. Specificity, or the true negative rate (TNR), is computed by dividing the number of correctly identified non-cancer episodes (“true negatives”) by the total number of the observed non-cancer episodes (“all negatives”). It measures the success rate of the classifier in correctly not recalling a client when cancer is absent in the client. This is a key performance metric because unnecessarily recalling clients to assessment center is costly and induces significant stress on clients and their families [44].

To compare the sensitivity and specificity of the AI-integrated screening scenarios with the current screening pathway, McNemar’s test (with continuity correction to improve the approximation of binomial distribution by the chi-squared distribution) is used to test for differences [45], and the binomial exact test (one-sided) is used to test for superiority. Both tests adhere to the correct design for McNemar’s test, in which a 2-by-2 contingency table is constructed based on the paired samples from the two comparison scenarios. The samples are paired by the episode ID, and the tests are conducted once for each of the sensitivity and specificity [46, 47] at a significance level of 5%.

## Notes

Funding: This work is supported by funding from the Australian Government under the Medical Research Future Fund Grant (MRFAI000090) for the Transforming Breast Cancer Screening with Artificial Intelligence (BRAIx) Project awarded to HMLF, DJM, PB, Jl, JH, GC and a National Health and Medical Research Council Investigator Grant (GNT1195595) awarded to DJM. This work is also supported by a Ramaciotti Health Investment Grant awarded to DJM and funding from a Royal Australian and New Zealand College of Radiologists Clinical Research Grant and the St Vincent’s Hospital Melbourne Research Endowment Fund awarded to HMLF. The funders had no role in the work or decision to publish.

### Funding Statement

This study was supported by funding from the Australian Government under the Medical Research Future Fund Grant (MRFAI000090) for the Transforming Breast Cancer Screening with Artificial Intelligence (BRAIx) Project and a National Health and Medical Research Council Investigator Grant (GNT1195595). This study was also supported by a Ramaciotti Health Investment Grant and funding from a Royal Australian and New Zealand College of Radiologists Clinical Research Grant and the St Vincent's Hospital Melbourne Research Endowment Fund. The funders had no role in the work or decision to publish.

### Author Declarations

Ethics Committee of St Vincents Hospital Melbourne gave ethical approval LNR/18/SVHM/162 for this work

### Summary of Updates

The main text had missing cross-reference numbers to Supplementary Figures and Supplementary Tables. These cross-references have been added in this revision.

