## Supplementary Material for "Comparison of AI-integrated pathways with human-AI interaction for population mammographic screening"

### Supplementary Figures

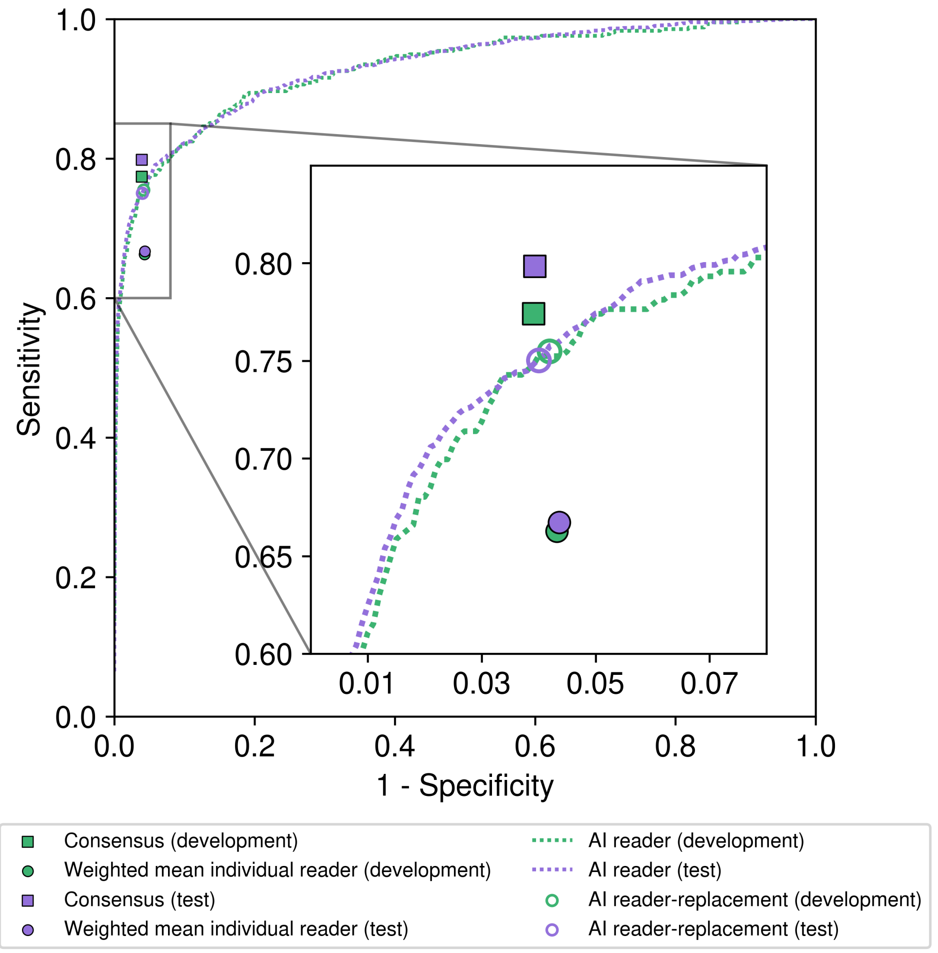


Appendix 1—figure 1. AI reader operating points on the development and test sets. The weighted mean individual reader performance is consistent over the development and test sets, leading to similar matched operating points on the respective ROC curves.

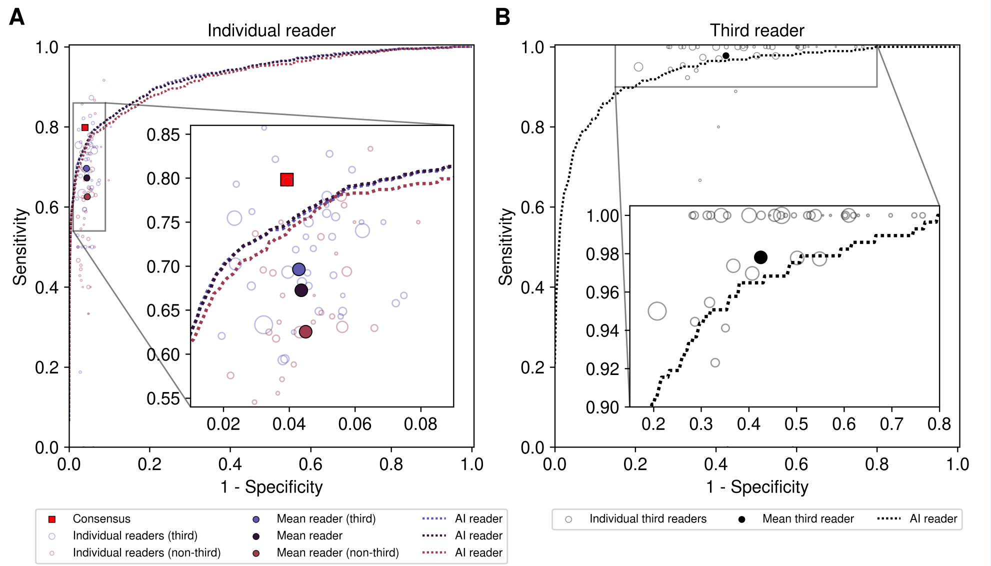


Appendix 1—figure 2. Performance of AI reader compared with reader groupings. (A) The AI reader ROC compared with mean individual readers grouped by whether or not they perform third reads in the dataset, individual readers also shown sized by number of reads. (B) The AI reader ROC compared with the mean third reader, individual third readers also shown sized by number of reads.

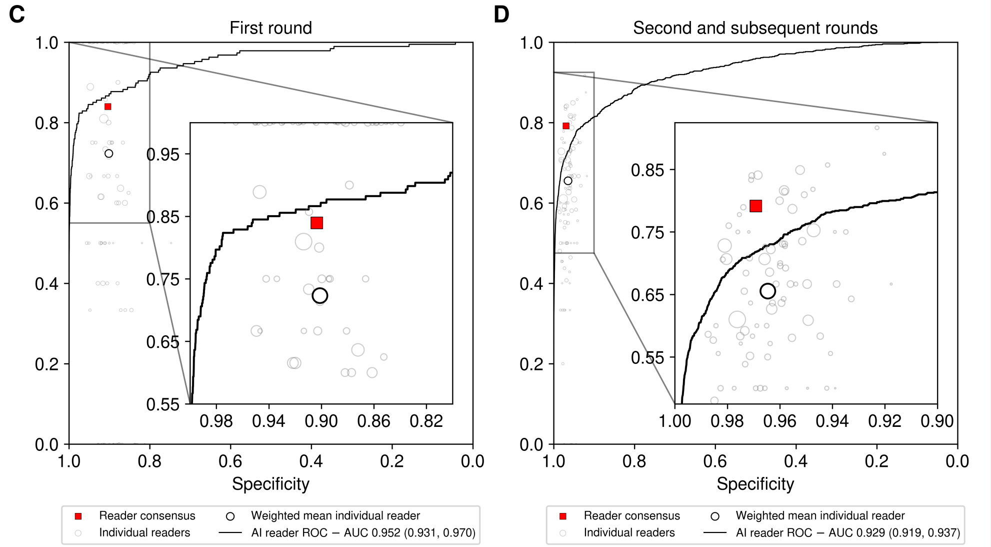


Appendix 1—figure 3. Performance of AI reader by round groupings. (A) The AI reader ROC compared with the weighted mean individual reader and reader consensus on first round episodes (n=19,374). Individual readers also shown sized by number of reads. (B) The AI reader ROC compared with the weighted mean individual reader and reader consensus on second and subsequent round episodes (n=129,731). Individual readers also shown sized by number of reads.

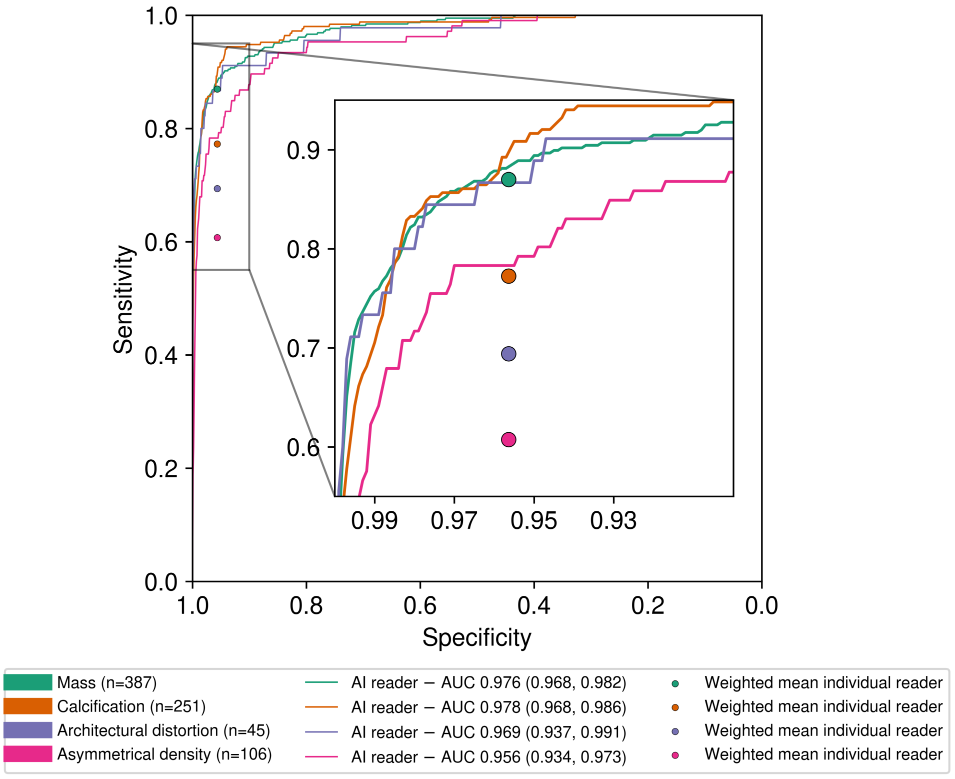


Appendix 1—figure 4. Performance of AI reader by radiologist morphology labels. The AI reader ROC compared with the weighted mean individual reader on groupings by morphology label. Screen-detected cancers only and when there was consensus agreement by both first and second reader or it went to a third reads, for each grouping all normal, NSA and benign are used as the negative class (n=147,776).

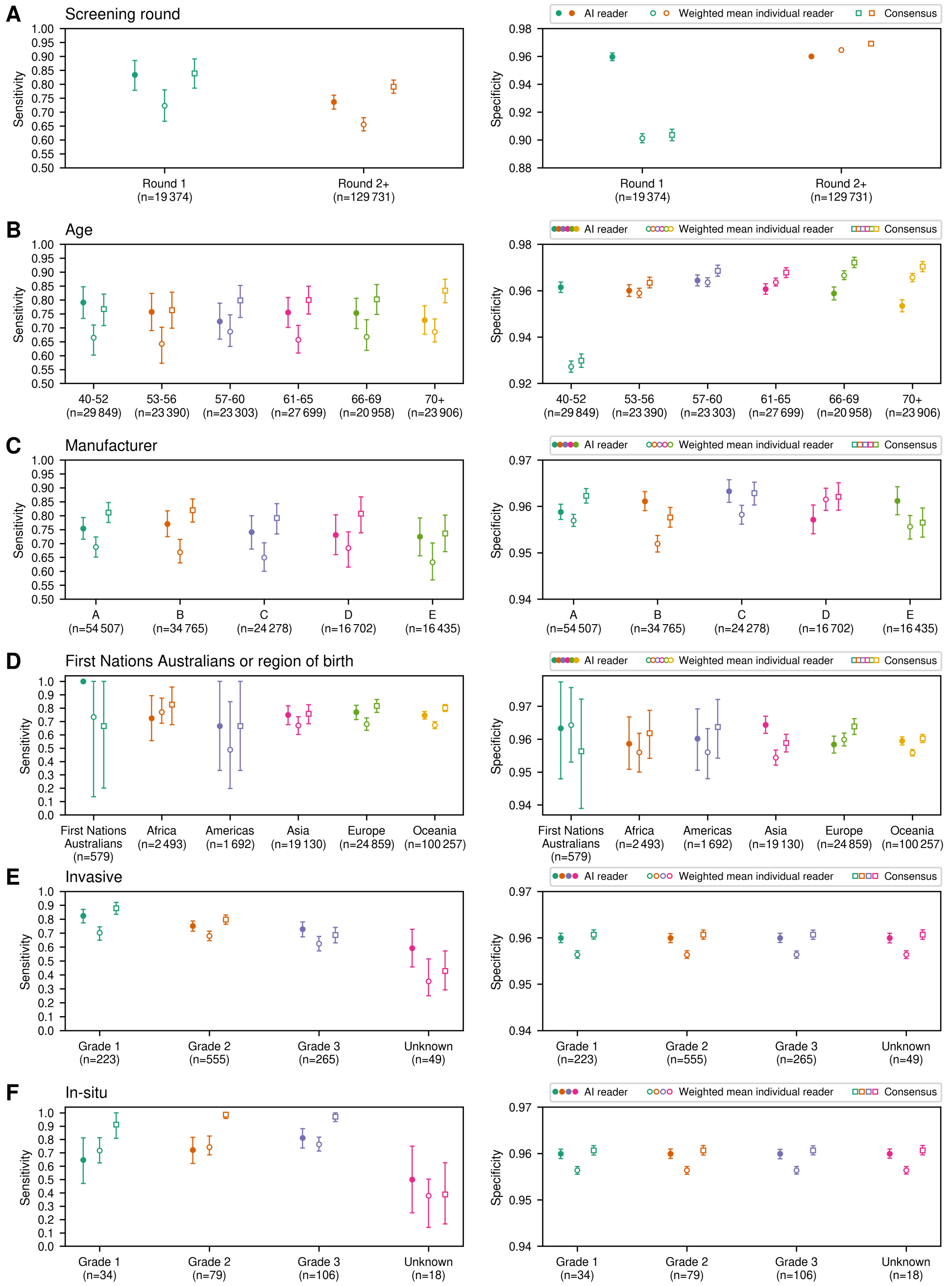

Appendix 1—figure 5. AI reader performance compared to radiologist performance on different episode groupings. The sensitivity and specificity with bootstrapped 95% confidence intervals of the AI reader at the AI reader-replacement operating points, the weighted mean individual reader and the consensus reader are reported for 2,000 replicates of each grouping. In D, the top three countries for each regional grouping were—Europe: England (23%), Italy (14%), Greece (10%); Asia: Vietnam (18%), China (16%), Philippines (9%); Americas: USA (30%), Chile (19%), Canada (14%); Africa: South Africa (35%), Egypt (20%), Mauritius (18%); and Australia & New Zealand: Australia (97%), New Zealand (2%), Fiji (< 1%).

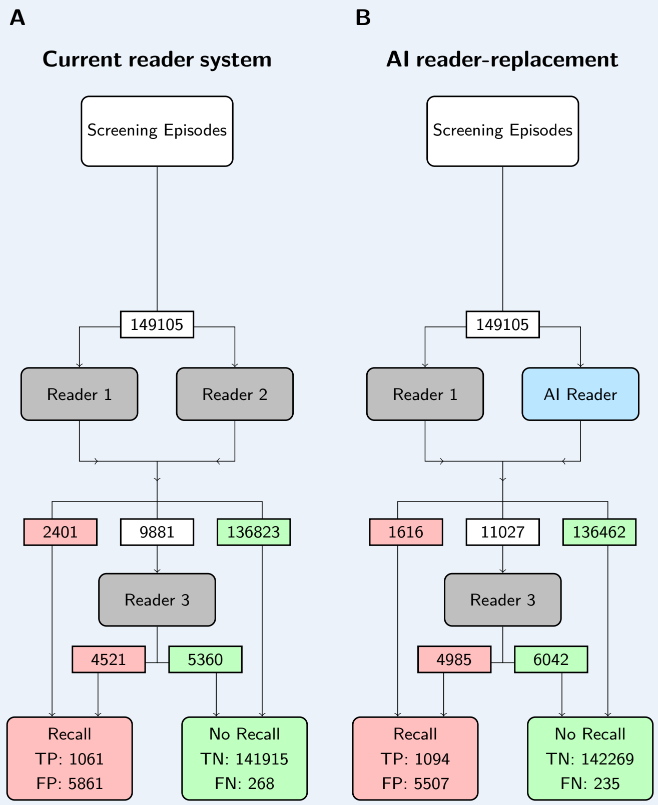


Appendix 1—figure 6. Screening episode flows for the current reader system and AI reader-replacement scenarios on retrospect test set. (A) The current reader system: Readers 1 and 2 see the same episode and opt to recall or not-recall, if they disagree Reader 3 arbitrates. Human readers perform 298,210 individual reads (Reader 1 and 2) and 9,881 arbitration reads (Reader 3). (B) AI reader-replacement: AI reader performs 50% (149,105) of total individual reads with an extra 1,146 human third reads required.

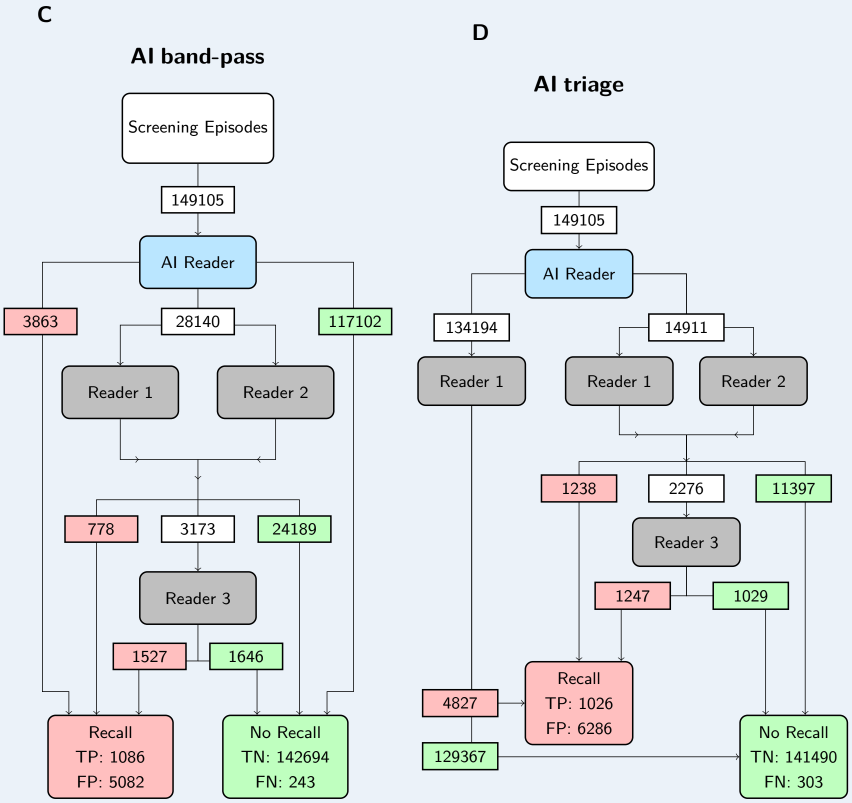


Appendix 1—figure 7. Screening episode flows for the AI band-pass and AI triage scenarios on retrospect test set. (C) AI band-pass scenario: AI reader screens out episodes before Readers 1 and 2. Episodes with high scores trigger the recall decision directly, and episodes with low scores trigger the no-recall decision directly. The other episodes continue to the usual screening pathway. AI reader performs 81.1% (120,965) of total individual reads with 6,708 fewer third reads required. (D) AI triage scenario: the AI reader triage's the episodes before Readers 1 and 2. Episodes with high scores continue to the usual pathway, and episodes with low scores go through the pathway with only 1 reader. 90% of episodes go through the 1-reader pathway, and overall 7,605 fewer third reads are required.

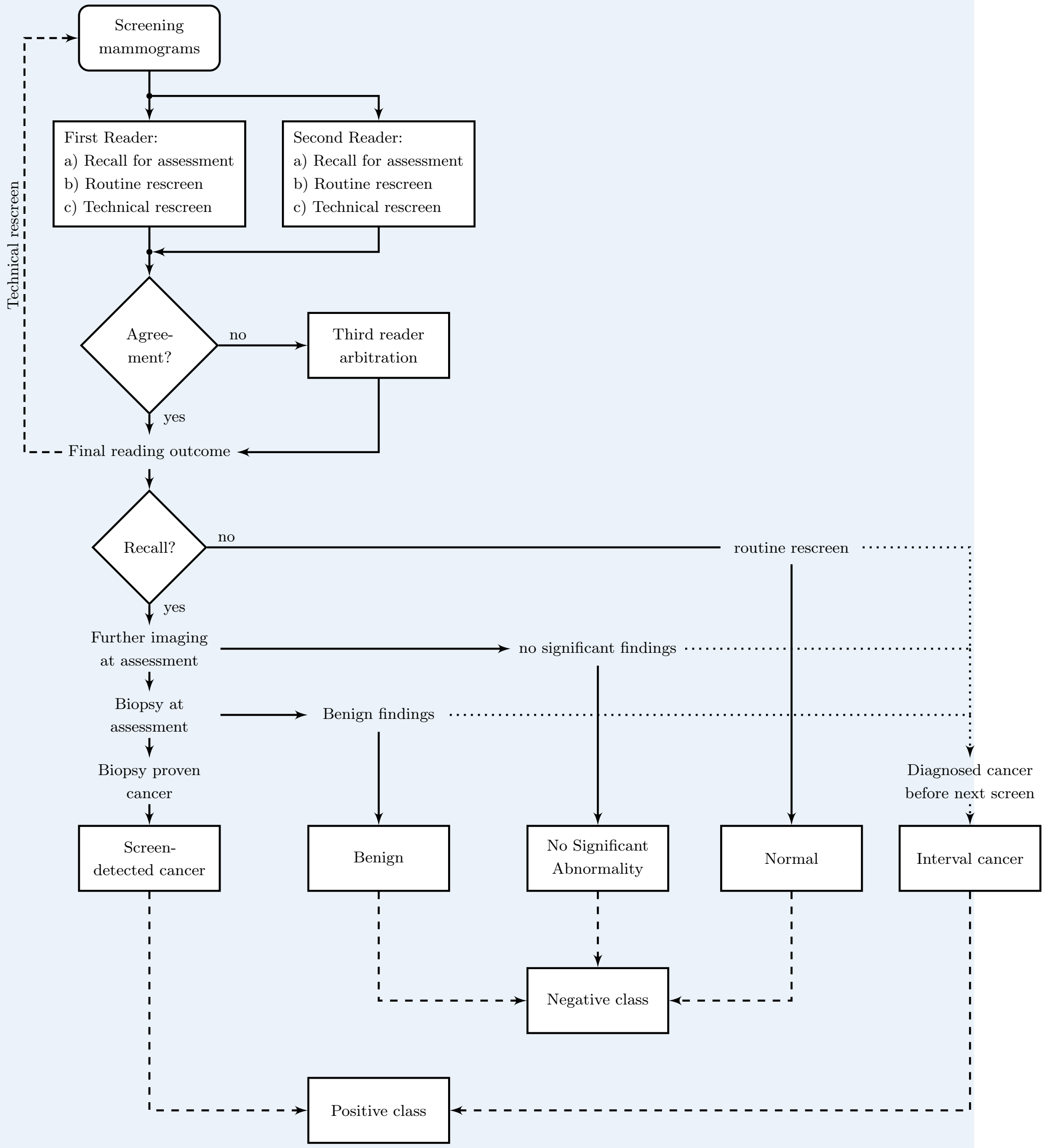


Appendix 1—figure 8. Flow chart of screening episode from mammogram acquisition to screening episode outcome.

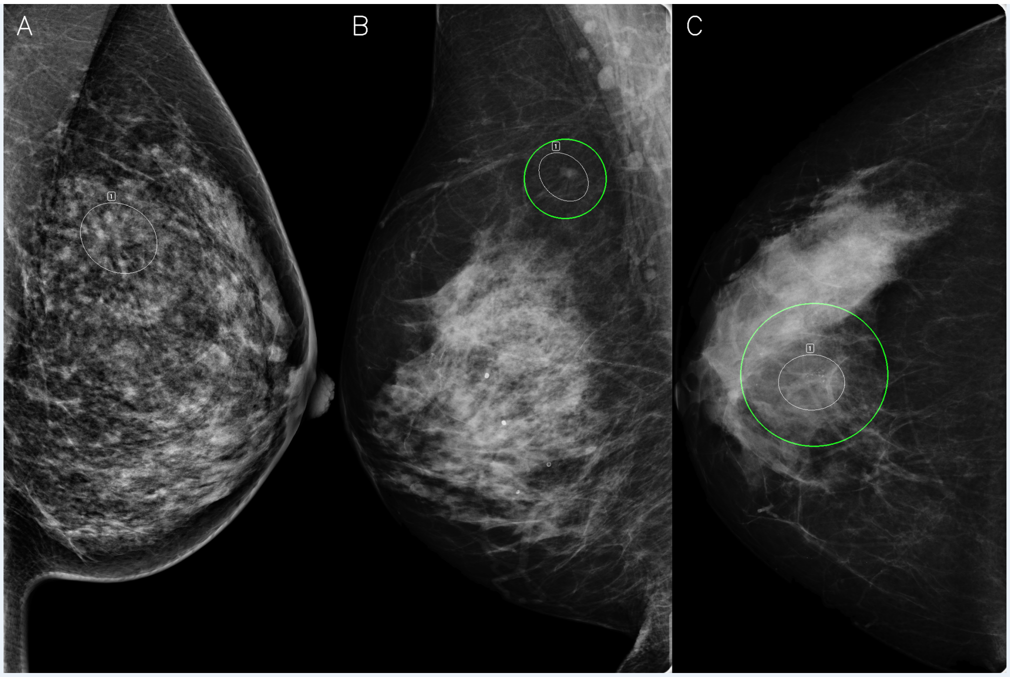


Appendix 1—figure 9. Three BRAIx AI reader annotation examples, BRAIX AI reader annotations are shown in white and original radiologist annotations in green (where available). (A) 45 year-old client with BI-RADS C density cleared at reading and subsequently presented with an interval cancer, invasive ductal carcinoma, in her left breast. The left medio-lateral oblique view demonstrates a subtle architectural distortion in the left upper breast identified by the BRAIx AI reader. Confirmation of agreement between BRAIx AI reader flagged area and exact site where interval cancer developed not possible as diagnostic imaging not available. (B) 67 year-old client, BI-RADS D density, with a spiculated mass in the upper right breast detected at screening. Ultrasound-guided core biopsy demonstrated invasive ductal carcinoma. (C) 56 year-old client, BI-RADS C density, with indeterminate microcalcifications in the central right breast detected at screening. Stereotactic core biopsy demonstrated ductal carcinoma in-situ.

### Supplementary Tables

| AI reader score bin | Screen-detected cancer | Interval cancer | Normal | Benign | NSA |
|---|---|---|---|---|---|
| 1 | 0 (0.0) | 4 (1.4) | 14,793 (10.4) | 30 (1.0) | 84 (3.0) |
| 2 | 1 (0.1) | 5 (1.7) | 14,688 (10.3) | 59 (2.0) | 157 (5.6) |
| 3 | 0 (0.0) | 11 (3.8) | 14,606 (10.3) | 95 (3.1) | 199 (7.1) |
| 4 | 2 (0.2) | 13 (4.5) | 14,513 (10.2) | 156 (5.2) | 226 (8.1) |
| 5 | 5 (0.5) | 13 (4.5) | 14,431 (10.2) | 189 (6.3) | 273 (9.8) |
| 6 | 6 (0.6) | 17 (5.9) | 14,369 (10.1) | 223 (7.4) | 295 (10.6) |
| 7 | 5 (0.5) | 26 (9.1) | 14,269 (10.1) | 293 (9.7) | 317 (11.4) |
| 8 | 21 (2.0) | 31 (10.8) | 14,116 (9.9) | 369 (12.2) | 374 (13.4) |
| 9 | 36 (3.5) | 46 (16.0) | 13,844 (9.8) | 565 (18.7) | 419 (15.0) |
| 10 | 966 (92.7) | 121 (42.2) | 12,339 (8.7) | 1039 (34.4) | 446 (16.0) |

Appendix 1—table 1. AI reader scores stratified by ground truth label. The AI reader scores binned by deciles. Bin 1 represents the episodes with the lowest 10% of AI reader scores and bin 10 the episodes with the highest 10%. The number of episodes for each category are shown for each bin, with the percentage of all episodes in the category falling into that bin shown in parentheses. The AI reader performed well on both screen-detected and interval cancers with the top 10% of AI reader scores containing 92.7% of screen-detected cancers, and 42.2% of interval cancers (81.8% of all cancers). The top 50% contained 99.2% of screen-detected cancers and 84.0% of interval cancers (95.9% of all cancers), while the bottom 50% contained 25.3% of all false positives (Benign and NSA), 0.8% of screen-detected cancers, and 15.9% of interval cancers.

| Dataset | Episodes | | | AUC | BRAIx | GMIC |
|---|---|---|---|---|---|---|
| ADMANI Prospective | Normal | 24990 | Breast | ROC | 0.976 (0.962, 0.989) | - |
| | Benign | 663 | | PR | 0.667 (0.601, 0.725) | - |
| | Screen-detected | 195 | Episode | ROC | 0.970 (0.953, 0.983) | - |
| | Interval | 0 | | PR | 0.670 (0.605, 0.728) | - |
| CSAW-CC[1] | Normal | 22868 | Breast | ROC | 0.994 (0.990, 0.997) | 0.943 (0.931, 0.954) |
| | Benign | 0 | | PR | 0.860 (0.832, 0.886) | 0.495 (0.447, 0.543) |
| | Screen-detected | 524 | Episode | ROC | 0.993 (0.991, 0.995) | - |
| | Interval | 0 | | PR | 0.889 (0.867, 0.909) | - |
| CSAW-CC | Normal | 23903 | Breast | ROC | 0.943 (0.933, 0.953) | - |
| | Benign | 0 | | PR | 0.651 (0.617, 0.682) | - |
| | Screen-detected | 524 | Episode | ROC | 0.934 (0.923, 0.944) | - |
| | Interval | 267 | | PR | 0.685 (0.655, 0.715) | - |
| CMMD | Normal | 0 | Breast | ROC | 0.906 (0.895, 0.918) | 0.831 (0.815, 0.846) |
| | Benign | 465 | | PR | 0.922 (0.911, 0.932) | 0.859 (0.842, 0.875) |
| | Screen-detected | 1310 | Episode | ROC | - | - |
| | Interval | 0 | | PR | - | - |
| BREAST | Normal | 361 | Breast | ROC | 0.972 (0.960, 0.982) | - |
| | Benign | 0 | | PR | 0.910 (0.876, 0.939) | - |
| | Screen-detected | 179 | Episode | ROC | 0.962 (0.947, 0.976) | - |
| | Interval | 0 | | PR | 0.939 (0.913, 0.961) | - |

Appendix 1—table 2. Results of testing the BRAIx AI readers on prospective and external datasets. 95% confidence interval were calculated using 2,000 bootstrap replicates. "Screen-detected cancer" and "Interval" cancer are abbreviated in the table to "Screen-detected" and "Interval", respectively. The three external datasets were: the Cohort of Screen-age Women - Case control (CSAW-CC) [48], the Chinese Mammography Dataset (CMMD) [49], and the BreastScreen Reader Assessment Strategy Australia (BREAST Australia) [50]. CSAW-CC is a subset of a large screening population dataset from Sweden. We report breast and episode level results for the entire dataset and screen-detected cancers only. CMMD is a publicly available, cancer enriched dataset from China. As not all clients in the CMMD dataset have four mammographic images we report breast level results only. BREAST is an Australian dataset of curated test sets designed to assess the performance of radiologists and radiology registrars. We present the breast and episode results for the combined test sets excluding any prior episodes. These datasets contain mammography vendors not used in the training of our models and no fine-tuning of the AI reader was done on these datasets. They were used purely as external test sets for the AI reader. Other datasets commonly used for comparisons - INbreast, DDSM, OPTIMAM - were considered, but omitted for reasons of sample size or licensing/access restrictions. These results improved significantly on previously reported results, represented here by the GMIC reader, which was the best-performing of six purported state-of-the-art AI readers in recent independent evaluations [51, 52].

[1] screen-detected cancers only

| | AI reader vs Radiologists | | | | | | | |
|---|---|---|---|---|---|---|---|---|
| Metric | T-T | T-F | F-T | F-F | McNemar test | P value | Binomial test | P value |
| Sensitivity | 801 | 196 | 103 | 229 | Significant difference | $1.035 \times 10^{-7}$ | AI superior | $4.105 \times 10^{-8}$ |
| Specificity | 136155 | 5702 | 5080 | 839 | Significant difference | $2.224 \times 10^{-9}$ | AI superior | $1.101 \times 10^{-9}$ |

Appendix 1—table 3. Testing the performance of the AI reader against the human reader on the retrospective dataset. The column T-F refers to the number of episodes for which AI reader made the correct assessments and the human reader made the incorrect assessments, and likewise for the T-T, F-T, and F-F columns. We use the McNemar test to test for differences, and we use the one-sided binomial exact test to establish superiority when the McNemar test rejects the null hypothesis. The AI reader shows superior sensitivity and specificity compared to the radiologists. Both screen-detected cancers and interval cancers are included.

| AI Standalone VS standard of care | | | | | | | | |
|---|---|---|---|---|---|---|---|---|
| Metric | T-T | T-F | F-T | F-F | McNemar test | P value | Binomial test | P value |
| Sensitivity | 191 | 77 | 141 | 920 | Significant difference | $1.982 \times 10^{-5}$ | AI inferior | $8.701 \times 10^{-6}$ |
| Specificity | 836 | 5024 | 5083 | 136833 | No difference | 0.564 | N/A | N/A |
| **AI Reader Replacement (RR) VS standard of care** | | | | | | | | |
| Metric | T-T | T-F | F-T | F-F | McNemar test | P value | Binomial test | P value |
| Sensitivity | 186 | 82 | 49 | 1012 | Significant difference | 0.00518 | RR superior | 0.00249 |
| Specificity | 2089 | 3772 | 3394 | 138521 | Significant difference | $8.448 \times 10^{-6}$ | RR superior | $4.205 \times 10^{-6}$ |
| **AI Band-Pass (BP) VS standard of care** | | | | | | | | |
| Metric | T-T | T-F | F-T | F-F | McNemar test | P value | Binomial test | P value |
| Sensitivity | 210 | 58 | 33 | 1028 | Significant difference | 0.0119 | BP superior | 0.00573 |
| Specificity | 2637 | 3224 | 2445 | 139470 | Significant difference | $4.997 \times 10^{-25}$ | BP superior | $2.111 \times 10^{-25}$ |
| **AI Triage (TR) VS standard of care** | | | | | | | | |
| Metric | T-T | T-F | F-T | F-F | McNemar test | P value | Binomial test | P value |
| Sensitivity | 266 | 2 | 37 | 1024 | Significant difference | $5.199 \times 10^{-8}$ | TR inferior | $1.421 \times 10^{-9}$ |
| Specificity | 4241 | 1620 | 2045 | 139870 | Significant difference | $2.493 \times 10^{-12}$ | TR inferior | $1.180 \times 10^{-12}$ |

Appendix 1—table 4. Testing the performance of the AI integrative scenarios against the standard of care on the retrospective dataset. The column T-F refers to the number of episodes that are correctly classified in the AI integrative scenario but incorrectly classified in the standard of care, and likewise for the T-T, F-T, and F-F columns. We use the McNemar test to test for differences, and we use the one-sided binomial exact test to establish superiority when the McNemar test rejects the null hypothesis. The AI reader replacement and AI band-pass scenarios have superior sensitivity and specificity compared with the current reader system, while the AI triage scenario has inferior sensitivity and specificity.

| No. of episodes | Test | All |
|---|---|---|
| Age group | | |
| 40 to 49 | 1,477 | 1,477 |
| 50 to 59 | 10,375 | 10,375 |
| 60 to 69 | 9,312 | 9,312 |
| 70 to 74 | 3,612 | 3,612 |
| 75+ | 1,072 | 1,072 |
| Screening round | | |
| 1 | 3,076 | 3,076 |
| 2+ | 22,772 | 22,772 |
| Manufacturer | | |
| A | 12,458 | 12,458 |
| B | 12,351 | 12,351 |
| Other | 1,039 | 1,039 |
| Country of birth | | |
| Australia | 16,851 | 16,851 |
| United Kingdom | 1,283 | 1,283 |
| Italy | 655 | 655 |
| China | 640 | 640 |
| Other | 6,419 | 6,419 |
| Risk category | | |
| None | 13 | 13 |
| Average | 22,963 | 22,963 |
| Moderate | 1,747 | 1,747 |
| High | 1,125 | 1,125 |
| Personal history of breast cancer | | |
| Yes | 219 | 219 |
| No | 25,629 | 25,629 |
| Recalled for assessment | 1,433 | 1,433 |
| Screening outcome | | |
| Screen-detected cancer | 195 | 195 |
| Interval cancer | 0 | 0 |
| Normal | 24,441 | 24,441 |
| Benign | 663 | 663 |
| No significant abnormality | 549 | 549 |
| Total | 25,848 | 25,848 |

Appendix 1—table 5. Summary and characteristics of prospective data.

| Reader 3 | Sensitivity (%) | Specificity (%) | Reader 3 / Total reads | Total cost |
|---|---|---|---|---|
| Fully simulated (original) | 82.3 (81.5, 83.1) | 96.3 (96.2, 96.3) | 11,027 / 160,132 | $10,456,696 |
| Second reader as third reader | 77.0 (76.3, 77.7) | 97.1 (97.1, 97.2) | 11,026 / 160,131 | $8,648,844 |
| Mixed simulated | 82.1 (81.4, 82.9) | 96.1 (96.1, 96.2) | 11,026 / 160,131 | $10,739,301 |

Appendix 1—table 6. Different ways to simulate Reader 3 in the AI reader replacement scenario. The fully simulated (original) third reader is based on the empirical performance of the real Reader 3. The second-reader-as-third-reader case uses the reads from the real Reader 3 when it is possible and otherwise from the reader that was replaced by the AI reader. Using reads from the replaced reader for arbitration ([19, 21]) is convenient, since the readers have seen all the episodes. However, this overlooks the fact that while Reader 1 and 2 make independent judgements, they are conditionally dependent. A difficult cancer case is difficult for any reader. In such cases, the two readers would frequently miss together, and the overall sensitivity would drop if either of them is used as an arbiter. In general, Reader 3 makes decisions differently than Readers 1 and 2 because Reader 3 has access to their decisions and analyses. Readers 1 and 2 should not be used in place of Reader 3 for simulation purposes. The mixed simulated third reader uses the reads from the real Reader 3 when it is possible and simulates otherwise. The mixed simulated case is consistent with the fully simulated but with reduced standard derivation because fewer simulations are required.
